# Genetic relationship between the immune system and autism spectrum disorder and traits

**DOI:** 10.1101/2023.10.16.23296896

**Authors:** Martina Arenella, Giuseppe Fanelli, Lambertus A. Kiemeney, Grainne McAlonan, Declan G. Murphy, Janita Bralten

**Affiliations:** Department of Forensic and Neurodevelopmental Science, Institute of Psychiatry, Psychology and Neuroscience, King’s College London, London, UK; Department of Human Genetics, Radboud university medical center, Nijmegen, the Netherlands; Donders Institute for Brain, Cognition and Behaviour, Nijmegen, The Netherlands; Department of Biomedical and Neuromotor Sciences, University of Bologna, Bologna, Italy; Department for Health Evidence, Radboud university medical center, Nijmegen, The Netherlands; Maudsley and South London NHS Foundation, London

**Author notes:** Corresponding author: Martina Arenella. Declaration of interest The authors declare no conflicts of interest.

**Keywords:** autism, autistic-like traits, autoimmunity, allergy, pleiotropy, GWAS, genetic correlation, polygenic risk, expression quantitative trait loci, methylation quantitative trait loci

## Abstract

Autism spectrum disorder (ASD) is a common and complex neurodevelopmental condition. The pathophysiology of ASD is poorly defined; however, it includes a strong genetic component and there is increasing evidence to support a role of immune dysregulation. Nonetheless, it is unclear which immune phenotypes link to ASD through genetics. Hence, we investigated the genetic correlation between ASD and diverse classes of immune conditions and markers; and if these immune-related genetic factors link to specific autistic-like traits in the population.

We estimated global and local genetic correlations between ASD (n=55,420) and 11 immune phenotypes (n=14,256–755,406) using genome-wide association study summary statistics. Subsequently, polygenic scores (PGS) for these immune phenotypes were calculated in a population-based sample (n=2,487) and associated to five autistic-like traits (i.e., attention to detail, childhood behaviour, imagination, rigidity, social skills), and a total autistic-like traits score. Sex-stratified PGS analyses were also performed.

At the genome-wide level, ASD was positively correlated with allergic diseases (ALG), and negatively correlated with lymphocyte count, rheumatoid arthritis (RA), and systemic lupus erythematosus (SLE) (FDR-p=0.01-0.02). At the local genetic level, ASD was correlated with RA, C-reactive protein, and granulocytes and lymphocyte counts (p=5.8x10-6–0.002). In the general population sample, increased genetic liability for SLE, RA, ALG, and lymphocyte levels, captured by PGS, was associated with the total autistic score and with rigidity and childhood behaviour (FDR-p=0.03).

In conclusion, we demonstrated a genetic relationship between ASD and immunity that depends on the type of immune phenotype considered; some increase likelihood whereas others may potentially help build resilience. Also, this relationship may be restricted to specific genetic loci and link to specific autistic dimensions (e.g., rigidity).

**Highlights:**

- Autism spectrum disorder (ASD) and diseases and traits linked to the immune system share genetic variability
- Positive genetic correlation is observed between ASD and atopic diseases
- Negative genetic correlation is observed between ASD and autoimmune diseases
- Shared genetic loci between ASD and immune-related phenotypes influence gene expression in immune-related tissues and in the developing brain
- Immunogenetic factors associate with rigidity and childhood behaviour in the population

## Introduction

Autism spectrum disorder (ASD) is a complex neurodevelopmental condition, with a strong genetic component and estimated heritability of 70-90% (Tick et al., 2016). ASD is common and it is diagnosed in approximately 1.6% of the population, with a 4:1 male-to-female ratio (Chiarotti & Venerosi, 2020). Clinically, ASD is characterised by different symptoms, that include impaired social communication and interaction abilities, repetitive patterns of behaviours and interests, and often atypical sensory processing (American Psychiatric Association, 2013). These symptoms generally persist throughout life and influence several aspects of personal and interpersonal functioning (van Heijst & Geurts, 2015). ASD symptoms also incur high social costs (medical and non) (Rogge & Janssen, 2019). Nevertheless, there is limited understanding of their underlying pathophysiological mechanisms.

**Figure 1.**
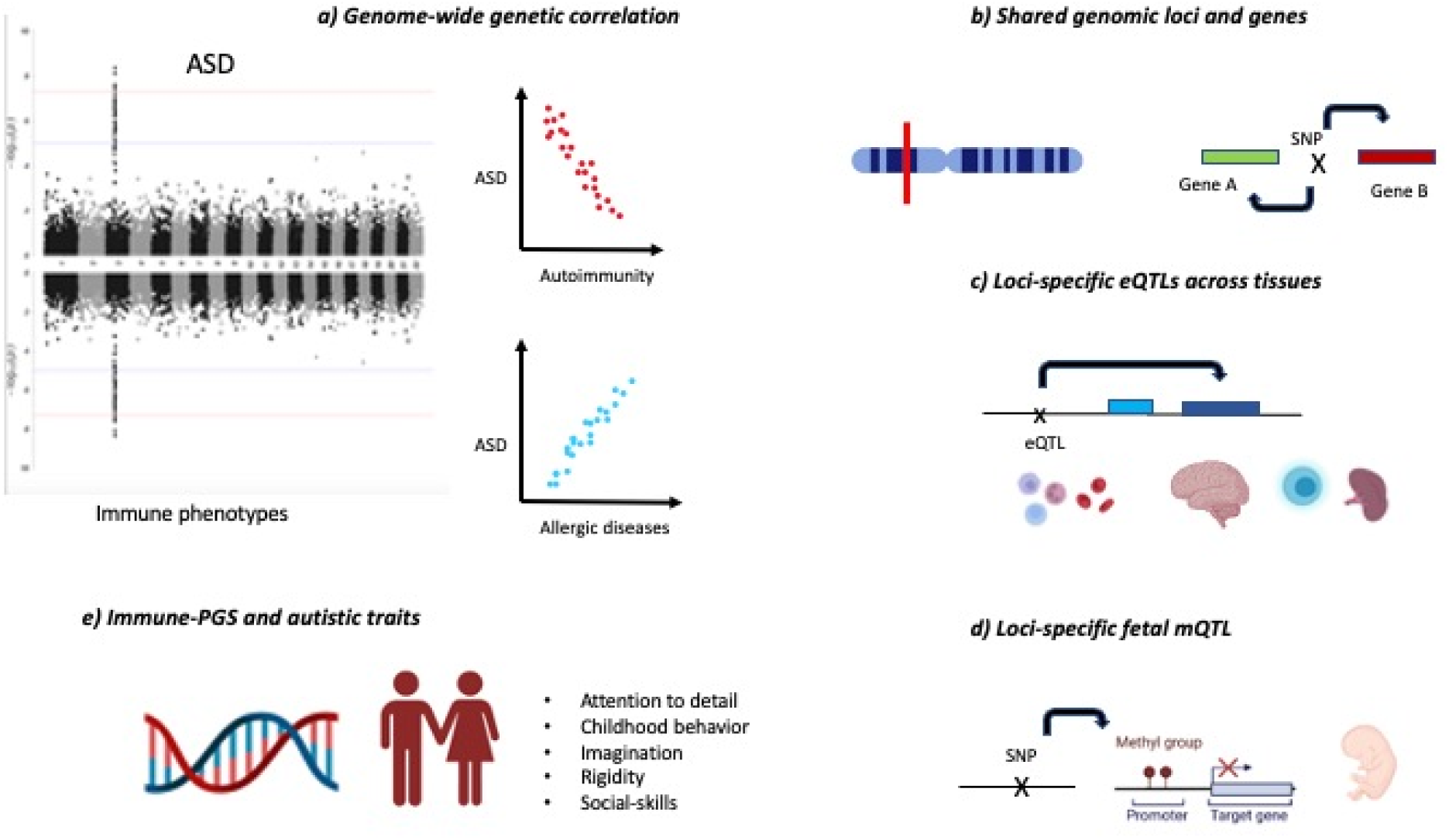
Illustrative graph of the analytical flow of the study. a) Genome-wide genetic correlation between ASD and immunity was estimated, using GWAS summary statistics for ASD and immune phenotypes. Both positive and negative genetic correlation were identified. b) Local genetic correlation analyses identified shared loci. Within the loci, SNPs were mapped to genes based on position. c) SNPs acting as eQTL were identified in each locus and mapped to genes expressed in brain and immune tissues. d) SNPs acting as mQTL in the fetal brain were identified in each locus and mapped to genes. e) In the general population, polygenic scores for immune phenotypes were associated with five autistic-like traits, and also across sexes

However, increasing evidence, from both animal and human studies, suggests that the immune system (and especially immune over-activation) may play a key role in ASD. For example, findings in rodents link maternal immune activation (MIA) during pregnancy to the onset of ASD-like behaviours in their offspring (Boulanger-Bertolus et al., 2018; Estes & McAllister, 2016; K. Liu et al., 2023). In humans, prior studies support the presence of inflammation and autoimmunity in autistic individuals, as indexed by increased blood levels of pro-inflammatory cytokines and anti-neuronal antibodies respectively (Edmiston et al., 2018; Mostafa et al., 2013). Also, there are reports of immune pathologies, like allergies, infections, and autoimmune diseases, in (a portion of) autistic individuals (Zerbo et al., 2015).

Notably, research in animals and in humans both suggest that the genetic factors may intervene in the relationship between ASD and immunity. For instance, experiments in animal models of MIA demonstrated that genetic regulators of immunity, such as interleukin- 17 pathway genes, may mediate the effects of MIA on the offspring behaviours. (G. B. Choi et al., 2016; Lombardo et al., 2018; Smith et al., 2007; Traglia et al., 2018). In humans, the contribution of immune genes to ASD is supported by i) epidemiological research which demonstrates an association between ASD and family history of autoimmune and inflammatory conditions (Atladóttir et al., 2009; Zerbo et al., 2015), and by ii) prior genetic studies. Namely, candidate gene analyses, reported an association between genes belonging to the human leukocyte antigen (HLA) region, such as *HLA-G, HLA-DRB1* and *HLA-DQB1* genes, and ASD (Bennabi et al., 2018; Torres et al., 2016). These genetic associations have been confirmed by hypotheses-free genetic approaches- such as genome-wide association studies (GWAS) that linked ASD and common genetic variants enriched in pathways controlling antigen presentation, and leukocyte and cytokine activation (Grove et al., 2019). Additionally, our group reported that common genetic variations in genes involved in inflammatory processes are related to specific autistic-like traits – such as rigidity and attention to detail – in the general population (Arenella et al., 2021). Transcriptomic studies further demonstrated that ASD is linked to dysregulated expression of immune genes. Specifically, mRNA analyses of post-mortem brain tissues in ASD demonstrated up- regulation of several immunoregulatory and inflammatory gene pathways (Gandal et al., 2018); and recent in vivo studies using magnetic resonance imaging ‘virtual histology’ approaches revealed that immune gene dysregulations characterise cortical regions where autistic individuals have anatomical variations from the neurotypical range (Ecker et al., 2022).

Taken together, these findings support a role of the immune system in the pathophysiology of ASD; and, in particular, suggest that immunogenetic factors are important. However, prior studies have implicated a wide range of immune mechanisms, from autoimmunity to inflammation (Ashwood & van de Water, 2004; McAllister, 2017), and it is unclear which particular immune phenotypes link to ASD through genetics. To address this challenge, we estimated the genetic correlation between ASD and diverse classes of immune conditions and markers. In addition, due to the heterogeneous phenotype of ASD we investigated if the immune-related genetic factors link to particular autistic-like traits in the population.

First, we tested the existence of genome-wide genetic correlations between different types of immune diseases or general markers of inflammation and clinically diagnosed ASD. As genetic correlation may not be constant throughout the genome, we subsequently explored local genetic correlations between these immune-related phenotypes and ASD. For the loci that were found to be significantly related we explored the role of loci-specific variants in immune regulation and brain development. Last, we investigated whether the aggregated genetic risk for immune diseases, as captured by polygenic scores, are associated with the severity of autistic-like traits in the general population. Additionally, given the sex differences in the prevalence of both ASD (Halladay et al., 2015) and immune conditions (Angum et al., 2020), we stratified the polygenic score analyses by sex.

## Materials and Methods

### Genome-wide association studies summary statistics

To explore the genetic relationships between ASD and immune-related phenotypes, we leveraged publicly available summary statistics of the largest genome-wide association studies (GWASs) on ASD and immune phenotypes (Table 1). Inclusion criteria for GWAS data were : European ancestry, annotation to the Genome Reference Consortium Human (GRCh) 37/hg19 build, and a sample size (N effective) > 5,000.

**Table 1.** Characteristics of the samples used as input for the genetic correlation and polygenic score (PGS) analyses.

| **Phenotype** | **N total** | **N cases** | **N controls** | **Reference** |
| --- | --- | --- | --- | --- |
| Systemic lupus erythematosus (SLE) | 14,256 | 5,201 | 9,066 | (Bentham et al., 2015) |
| Rheumatoid arthritis (RA) | 58,284 | 14,361 | 43,923 | (Okada et al., 2014) |
| Autoimmune thyroid disease (AIT) | 755,406 | 30,234 | 725,172 | (Saevarsdottir et al., 2020) |
| Type 1 diabetes mellitus (T1DM) | 24,840 | 9,358 | 15,705 | (Forgetta et al., 2020) |
| Asthma | 303,859 | 64,538 | 239,321 | (Y. Han et al., 2020) |
| Allergic disease (ALG) | 102,453 | 25,685 | 76,768 | (Zhu et al., 2018) |
| Celiac disease (CD) | 15,283 | 4,533 | 10,750 | (Dubois et al., 2010) |
| Autism spectrum disorder (ASD) | 55,420 | 22,458 | 29,386 | (Matoba et al., 2020) |
| Lymphocyte count (LYMPH) | 408,112 | - | - | (Vuckovic et al., 2020) |
| Lymphocyte percentage (LYMP%) | 408,112 | - | - | (Vuckovic et al., 2020) |
| Neutrophil count<br>(NEU) | 408,112 | - | - | (Vuckovic et al., 2020) |
| Monocyte count<br>(MON) | 408,112 | - | - | (Vuckovic et al., 2020) |
| Eosinophil count<br>(EOS) | 408,112 | - | - | (Vuckovic et al., 2020) |
| C-reactive protein<br>(CRP) | 401,696 | - | - | (X. Han et al., 2020) |

### Autism spectrum disorders

We used the summary statistics of the meta-analysis of the GWAS of ASD including seven cohorts, from the iPSYCH, Psychiatric Genomic Consortium samples, and the Simon Foundation Powering Autism Research for Knowledge (SPARK) sample (N = 55,420) (Matoba et al., 2020).

### Immune phenotypes

We used GWAS summary statistics for;

a. a set of immune-related diseases that have been reported in autistic individuals and their families (Atladóttir et al., 2009; Zerbo et al., 2015), including autoimmune thyroid diseases (AID) (Saevarsdottir et al., 2020), celiac disease (CD)(Dubois et al., 2010), rheumatoid arthritis (RA) (Okada et al., 2014), systemic lupus erythematosus (SLE) (Bentham et al., 2015), and type 1 diabetes mellitus (T1DM) (Forgetta et al., 2020), and conditions associated with hypersensitivity to allergens (i.e., allergic diseases (ALG) (Zhu et al., 2018) and asthma (Y. Han et al., 2020));
b. blood levels of C-reactive protein (CRP) (X. Han et al., 2020), a peripheral biomarker of inflammation; and
c. the total counts (and/or relative percentage) of white blood cells (Vuckovic et al., 2020) involved in the fast response to infection (neutrophils), T and B cell-mediated response (lymphocytes), allergic reaction (eosinophils), and phagocytosis (monocytes).

### Genotype data for autistic-like traits

To examine whether the immune-related phenotypes that are genetically correlated with ASD, are linked to specific autistic dimensions in the general population, we explored population-based genotype data and measures of autistic-like traits in the Nijmegen Biomedical Study.

### The Nijmegen Biomedical Study (NBS)

We used genotype and behavioural data from a Dutch population-based cohort of 2,847 individuals who participated in the Nijmegen Biomedical Study (NBS) (mean age 28.4; 54% females). The NBS is a project managed by the Department for Health Evidence and the Department of Laboratory Medicine of the Radboud university medical center. The study was approved by the Institutional Review Board (CMO 2001/055) and aimed to investigate genetic factors, lifestyle, and environmental exposures underlying a range of traits and diseases (for further information, see (Galesloot et al., 2017)). In this cohort, genotyping was performed using the Illumina Human OmniExpress Beadchip platform. Initial single nucleotide polymorphism (SNP) filtering was applied on call rate (>95%), Hardy-Weinberg equilibrium (HWE<1x10^-6^), minor allele frequency (MAF > 0.01), and imputation quality (> 0.7). Autosomal SNPs were imputed to the 1000 Genome Reference Panel (1KGRP) phase 3 release, using Minimach. To assess population structure, multidimensional scaling (MDS) was performed in PLINK (Purcell et al., 2007) and the first four MDS components were retained as covariates in subsequent analyses. In addition, participants were asked to complete a self-report questionnaire on autistic-like traits, developed by qualified clinicians at Radboudumc and previously validated in the Dutch population (Arenella et al., 2022; Bralten et al., 2018). The questionnaire consists of 18-items, rated on a 4-point Likert scale, based on the autism quotient (AQ) questionnaire and the ASD criteria listed in the Diagnostic and Statistical Manual of Mental disorder – 5^th^ edition. The items cover the three main dimensions of ASD (social communication, social interaction, and repetitive behaviours). Moreover, some items enquire about the level of autistic behaviours (based on the DSM) present in childhood (i.e., childhood behavior). Factor analysis of these items identified five autistic-like traits: attention-to-detail, imagination, rigidity, social skills, and childhood behaviour (see Table S1, and (Arenella et al., 2022; Bralten et al., 2018) further details). These traits, along with a total autistic-like traits score, were normalised and adopted as target phenotypes for polygenic risk score (PGS) analyses described below.

### Shared genetic etiology between ASD and immune phenotypes

#### Global genetic correlations analyses

Global genetic correlation was estimated between ASD and immune-related diseases (i.e., AIT, ALG, Asthma, CD, RA, SLE, T1DM), and population-based variations in immune-inflammatory response as indexed by CRP blood levels, the blood count of eosinophils, lymphocytes, monocytes, and neutrophils (Table 1). Pair-wise global genetic correlation between ASD and each immune-related phenotype was estimated via Linkage Disequilibrium SCore (LDSC) regression as implemented in the LDSC v1.0.1 tool (https://github.com/bulik/ldsc) (Bulik-Sullivan et al., 2015). Analyses used pre-computed linkage disequilibrium (LD) scores based on the 1kGRP reference, which are suitable for European-centred GWASs. LDSC analyses consisted of two steps: 1) converting summary statistics data to the standard LDSC format (i.e., exclusion of HLA region and merging to the HapMap3 reference panel); 2) estimating genetic correlation. A block jack-knife procedure was used to estimate standard errors and calculate corresponding *p*-values. P-values of genetic correlation estimates (*rg*) were false discovery rate (FDR)-corrected, given the medium-high genetic intercorrelations and co-heritability of the immune phenotypes themselves (Fig 1; Table S2). Global genetic correlation analyses were restricted to GWAS summary statistics with sample size > 5,000 individuals, SNP-based heritability (h^2^_SNP_) > 0.05 and mean chi square > 1.02 as recommended in (Zheng et al., 2017).

**Figure 1.**
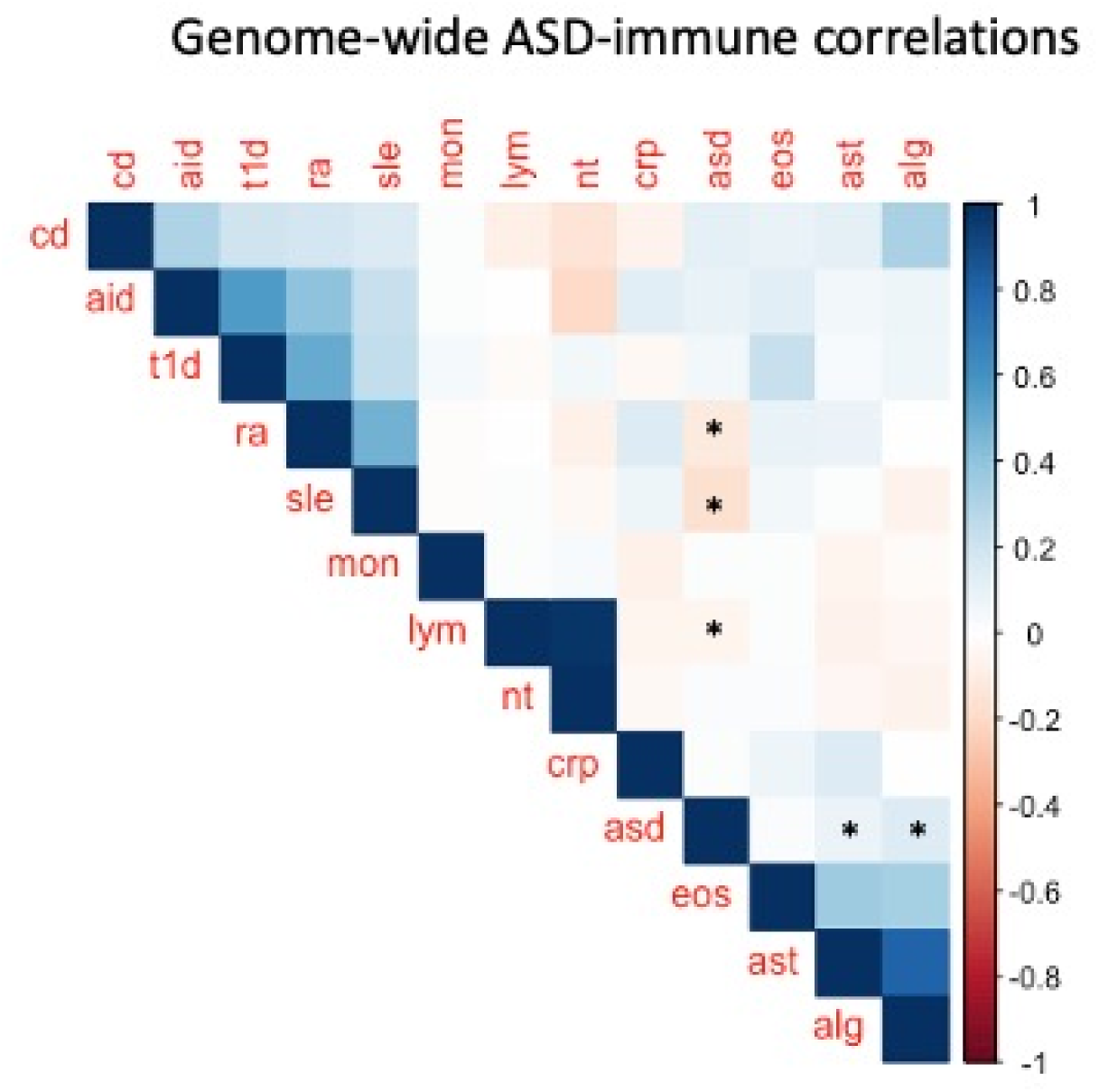
Genetic correlation plot summarising the results of the global genetic correlation analyses between ASD and immune-related phenotypes. Colour bar indicates variation in the strength and direction of genetic correlation estimates (rg) with positive rg in blue and negative rg in red. The FDR-corrected significant correlations are marked with an asterisk (*).

**Figure 2.**
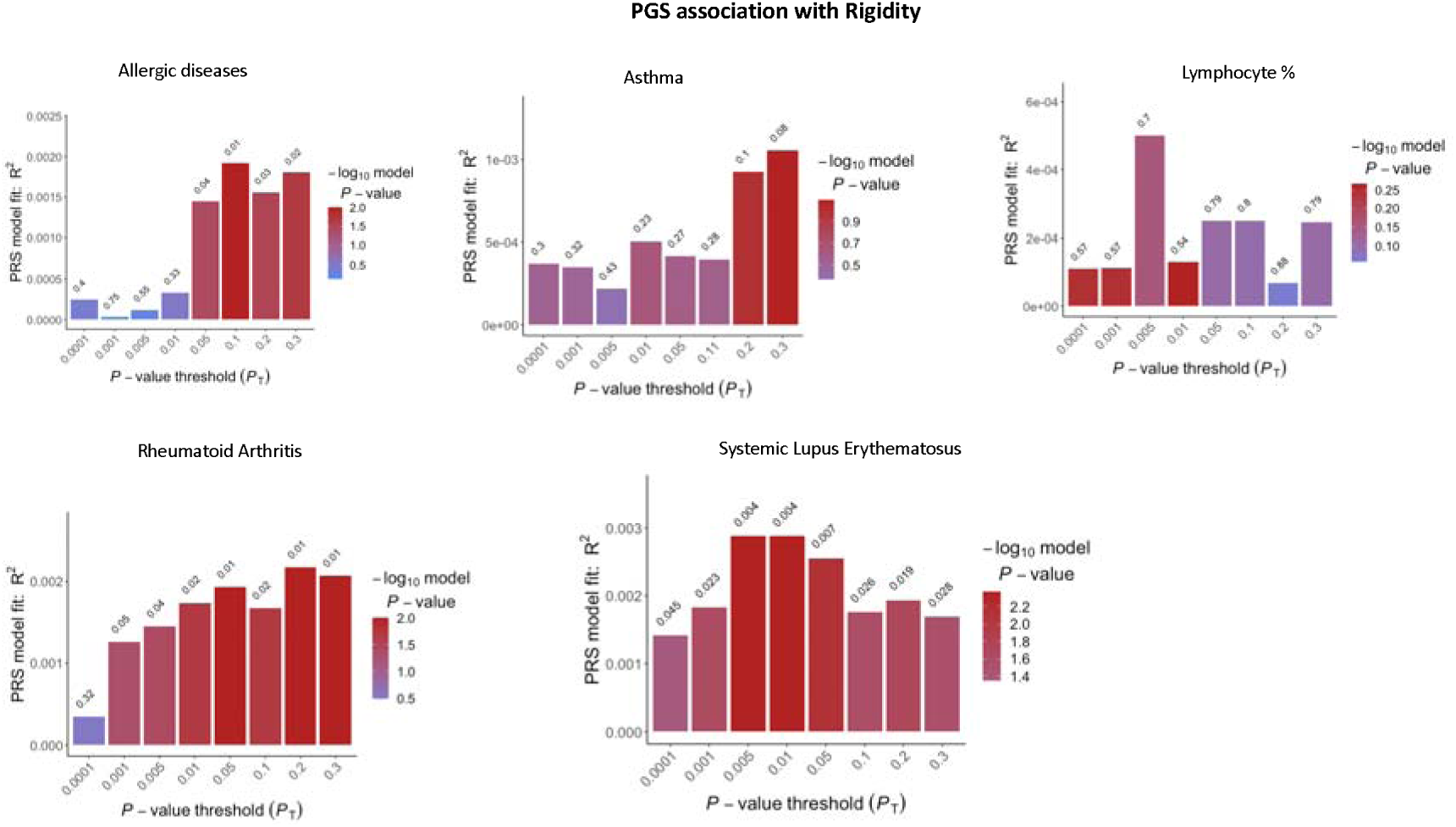
Bar plots for the association between polygenic scores for different immune-related phenotypes and rigidity. Each bar corresponds to the PGS calculated at the GWAS p-value threshold (Pt) listed on the x-axis. The height of the bar (y-axis) represents the degree of variance explained by each PGS in rigidity. The bar colour indicates the significance of the association (according to the -log10(p-value)). The p-value of association for each PGS is reported on the top of each bar.

### Local genetic correlation analyses

Local genetic correlation analyses complemented global genetic correlation between ASD and the immune-related phenotypes. This step allowed us to identify scenarios in which ASD-immune genetic correlations are restricted to specific genomic regions, and to determine shared genetic factors located in the genomic regions. Local genetic correlation analyses were performed using the R-package ‘Local Analysis of [co]Variant Association (LAVA)’ (Werme et al., 2022). Local genetic correlation was estimated across 2,495 loci defined by partitioning the genome into blocks of ∼1 Mb while minimising LD between them. The analyses consisted of two steps: 1) univariate association analyses, to detect the local h^2^_SNP_ signal of each phenotype within each genomic locus; and 2) bivariate association analyses, to estimate the pair-wise genetic correlation between two phenotypes of interest at the chosen locus. Bivariate association analyses were restricted to those genomic loci showing a significant h^2^ signal for both phenotypes (p<1x10^-4^). The p-values of local *rg* were Bonferroni-corrected, considering the number of loci tested in the bivariate association analyses. Subsequently, we identified SNPs included in each locus based on GRCh 37 positions and mapped these SNPs to genes using the *g:snpense* function of the R-package ‘g:profiler2’ (Kull et al., 2007). In addition, we queried to PubMed to explored if identified genes have been implicated in immunity. Since some SNPs (i.e., *cis-*eQTLs) may influence the transcription of proximal genes with differences across tissues, we tested if SNPs within each locus were linked to gene expression across tissues. For this, we used ‘e-MAGMA’ (https://github.com/eskederks/eMAGMA-tutorial) (Gerring et al., 2021), a tool which converts GWAS summary statistics for a phenotype of interest into a e-gene-level statistics that refers to a gene expressed in a given tissue (e-gene). The conversion takes into account LD between SNPs, and is based on a reference list of eQTL-to-gene association (FDR p- value<0.05) across different tissues from GTEx v8 (https://www.gtexportal.org/). The considered tissues were those relevant to neurodevelopment (brain cortex) and immune regulation and activation (spleen, lymphocytes, and whole blood).

To further investigate the impact of SNPs mapped within each genetically correlated locus on gene regulation, we examined their influence on *cis-*methylation of CpG sites in the developing brain (i.e., if they act as fetal mQTLs). To explore this, we adopted a frequentist approach and tested the enrichment of fetal mQTLs among loci-specific SNPs (Fisher’s exact test) (van Belle et al., 2004). This analysis was based on a publicly available compendium of ∼16,000 Bonferroni significant mQTLs in the developing brain (see (Hannon et al., 2016); https://epigenetics.essex.ac.uk/mQTL/).

### Polygenic score analyses

Polygenic score (PGS) analyses were conducted to explore if the additive effect of common genetic variants to the immune-related phenotypes was associated with autistic-like traits in a population-based sample. Linear regression models were used to test the association of the genetic liability to immune-related phenotypes with five autistic-like traits (i.e., rigidity, attention-to-detail, social skills, imagination, childhood behaviour) and the total autistic score in the target NBS cohort.

The GWAS summary statistics for the immune-related phenotypes showing genome-wide genetic correlation with ASD were individually used as base datasets for PGS calculation. The summary statistics underwent a preliminary clumping step using PLINK (Purcell et al., 2007) to ensure that only the most significant independent SNP for each LD block (r2>0.25, clumping window=±500kb) was considered. PGSs were then calculated on the target NBS individual-level genotype data using PRSice2 (S. W. Choi & O’Reilly, 2019). For each base immune phenotype showing genome-wide genetic correlation with ASD, PGSs were computed including SNPs exceeding seven *a priori* defined GWAS *p*-value thresholds (Pt) (i.e., Pt=0.0001, 0.001, 0.01, 0.05, 0.1, 0.2, 0.3). Multiple linear regressions were performed, considering PGSs for the immune-related phenotypes as independent variables and autistic-like traits as dependent variables. Age, sex, body mass index (BMI), and population structure (MDS) components were included as covariates. The variance explained by the PGS of each immune-related phenotype (PRS-R2 = full model R2 – null model R2) for each of the five autistic-like traits and the total score was calculated separately. The p-values of each association test were adjusted using FDR correction, considering the number of autistic-like traits and immune-related phenotypes tested. Results were considered statistically significant when p_FDR_ <0.05.

### Sex-stratified PGS analyses

To investigate if associations between immune-based PGS and autistic-like traits were sex-specific, PGS analyses were performed after stratifying the target NBS sample according to sex. Hence, we tested multiple general linear models for both sex group which included age, BMI, and MDS components as covariates. To reduce the burden of multiple between-sexes comparisons, and also minimise variations in effect size, we considered for each immune disease, the PGS that best explain variability in each autistic trait, so-called ‘best-fit’ PGS. We considered results statistically significant only if p_FDR_ <0.05.

## Results

### Global genetic correlations between ASD and immune phenotypes

We identified significant positive global genetic correlations between ASD and asthma (*rg*=0.08, se=0.006, p_FDR_= 0.02) and between ASD and allergic diseases (*rg*=0.14, se=0.1, p_FDR_=0.01). Additionally, ASD showed significant negative genetic correlations with autoimmune disorders (RA and SLE) and lymphocyte count/percentage (rg = -0.06-0.17; se = 0.02-0.06; p_FDR_=0.01) (Figure 1, Table S3).

### Local genetic correlations between ASD and immune phenotypes

For each ASD-immune pairwise comparison, we identified multiple loci with significant h^2^_SNP_ for both ASD and the immune phenotype considered (p < 1x10-4) (see Table S3). Of these loci, we registered significant genetic correlation – i.e., surviving multiple comparison correction – at 11 unique loci shared between ASD and AIT, RA, CRP, EOS, LYMP, MON, NEU. Among those, two loci - the chr11:95-96Mb locus and the chr17: 43-44Mb locus – showed genetic correlation between ASD and multiple immune phenotypes (AIT, EOS, and Lymph). We also observed local genetic correlation between ASD and CRP at the chr6:29- 30Mb locus containing the HLA region, a key immune-related region that is not covered by the LDSC analyses. Table 2 illustrates the significant genetic correlation loci and the genes belonging to these genetic regions.

**Table 2.**
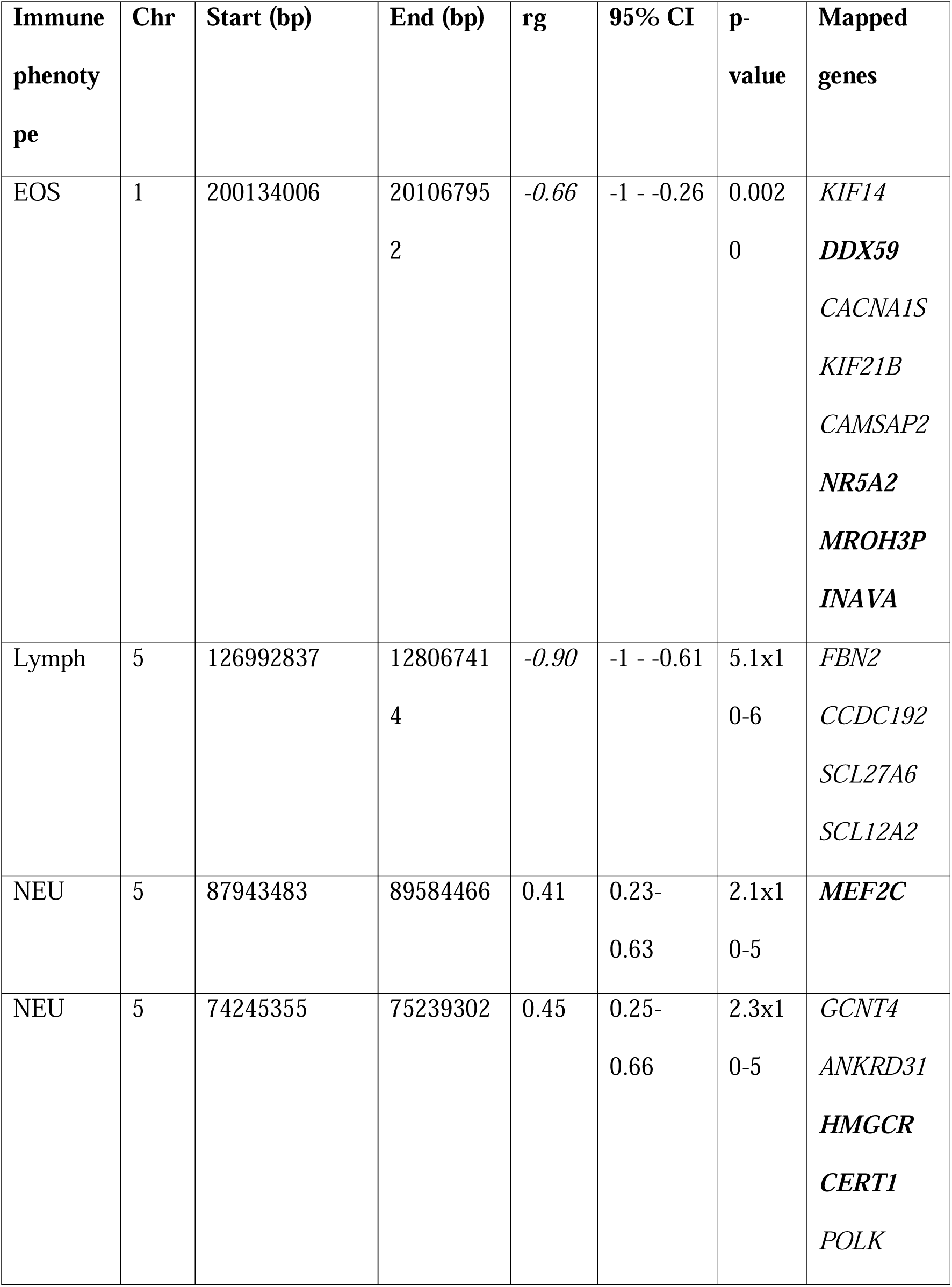
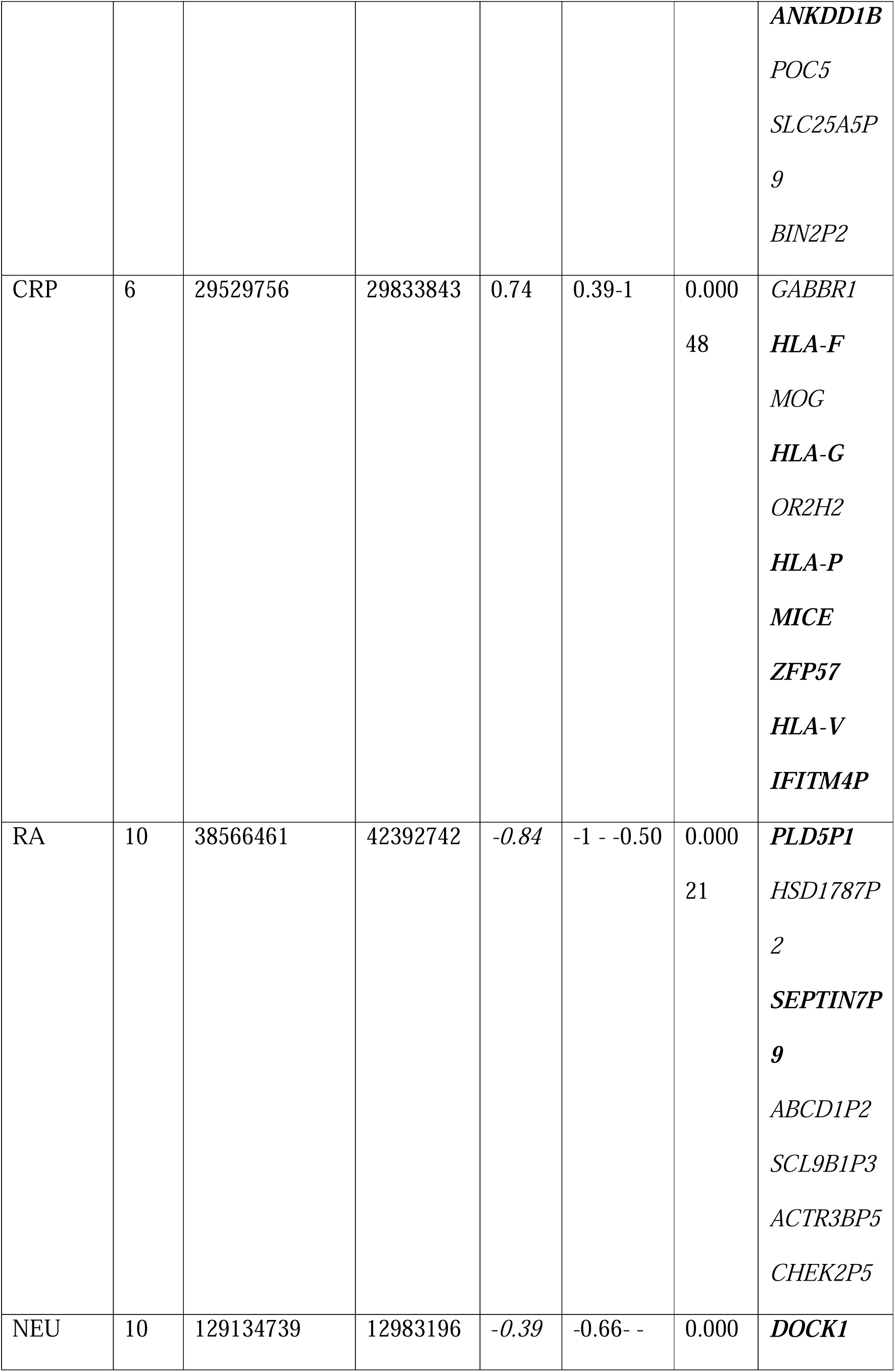
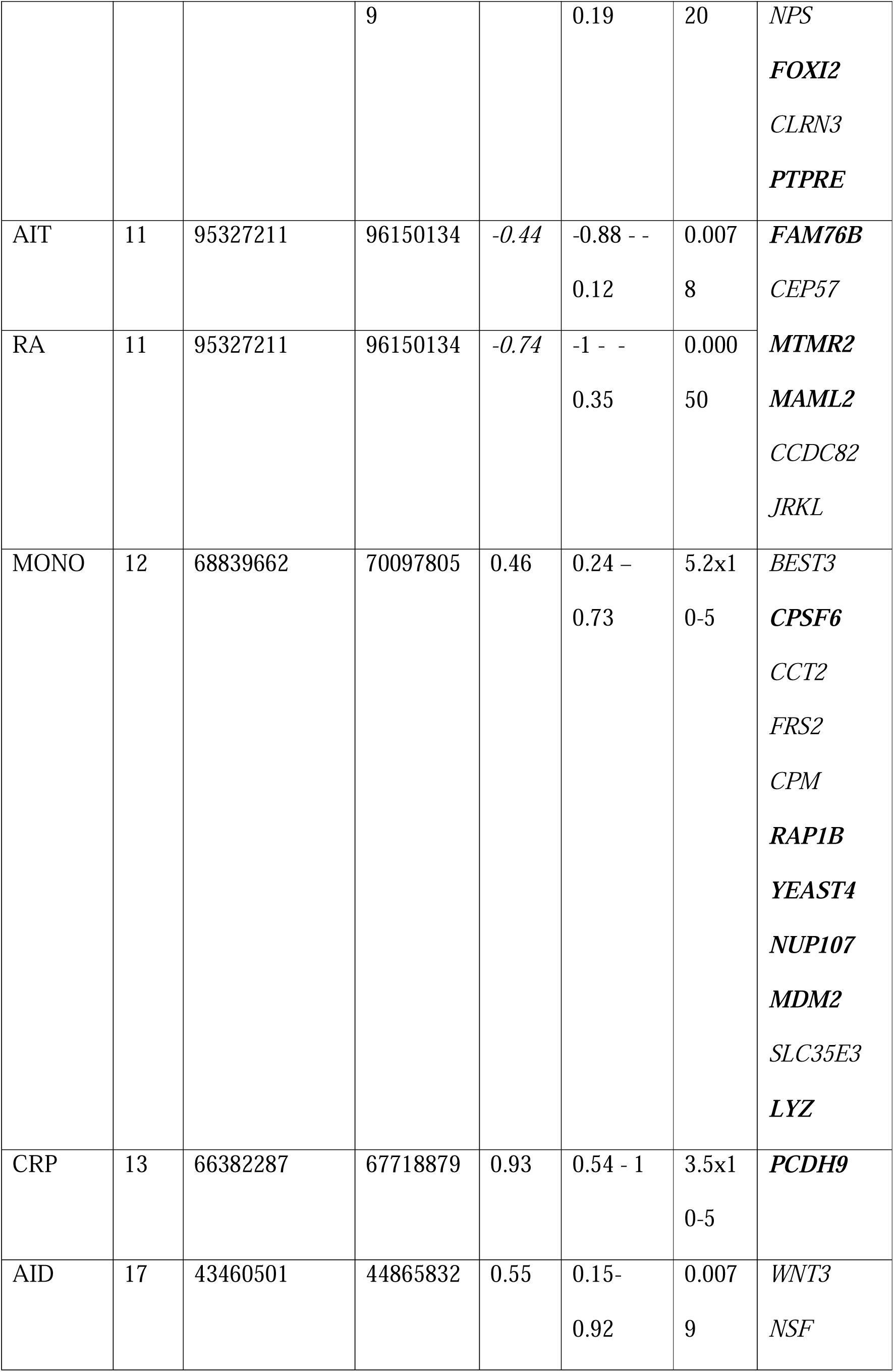
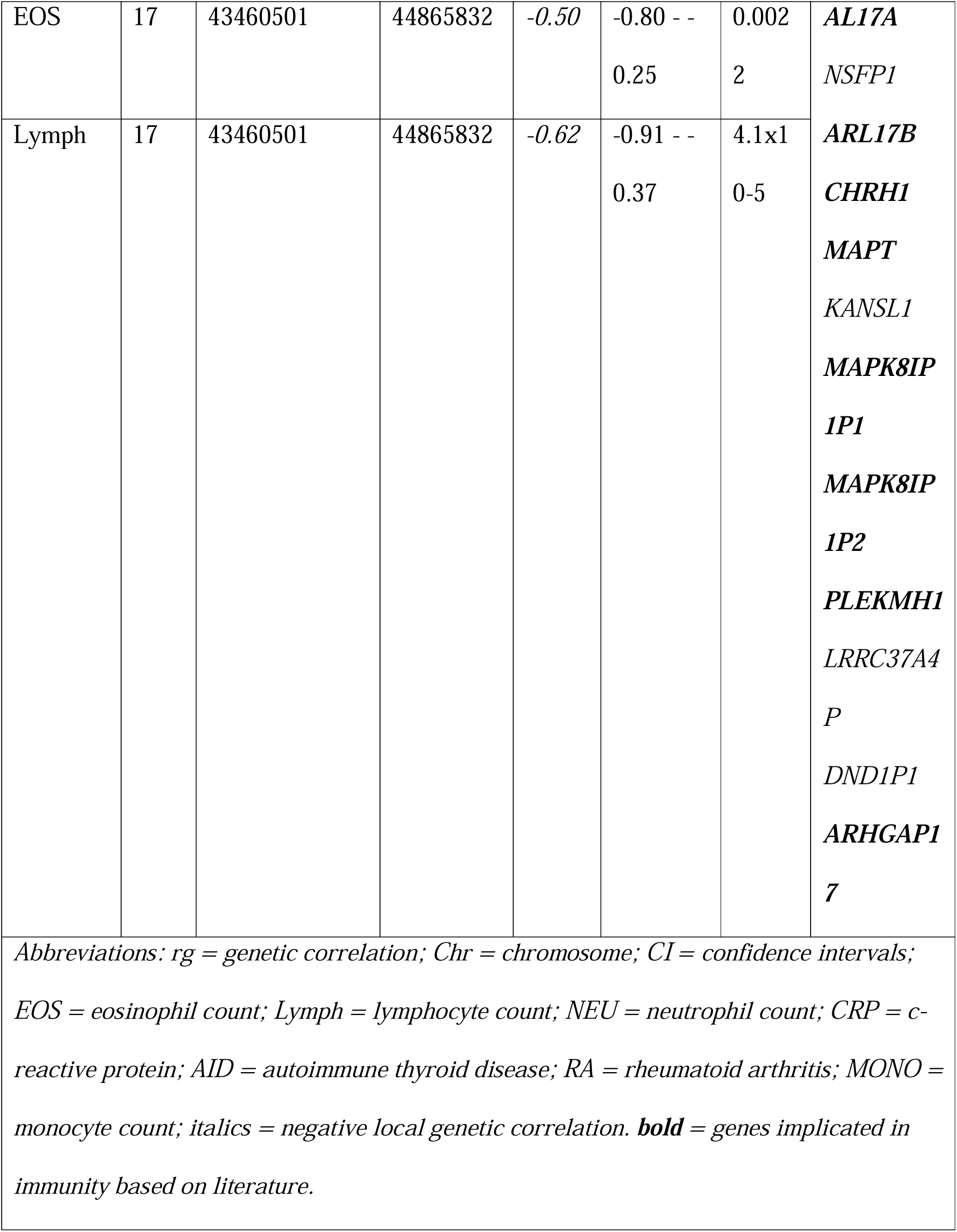
Loci with a Bonferroni-significant genetic correlation signal between ASD and immune-related phenotypes.

### Brain and immune-related eQTLs in shared loci

For four out of the 11 shared genomic loci between ASD and immune-related phenotypes (chr1:200-201Mb, chr6:29-30Mb, chr12:68-70Mb, chr17:43-44Mb), we identified e-genes, expressed in the brain and in immune tissues (Table S4). Specifically, at the chr1:200-201Mb locus, *DDX9* expression in the cortex and immune cells was significantly associated with ASD and eosinophil count (p = 0.01-0.0009); at the locus chr6:29-30Mb, the expression of *RNF39* in the cortex and the expression of *HLA-F* and *ZFP57* immune tissues were significantly associated with ASD and CRP (p = 0.03-2.9x10-5); at the locus chr12:68-70Mb, the expression of *YEATS4* in the cortex and immune cells, and the expression of *LYZ* and *MDM2* was significantly associated with ASD and monocyte count (p= 0.01-3.1x10-8); and last, at the locus chr17:43-44Mb, the expression of *KANSL1, ARL17A, LRRC37A2, LRRC37A* in the brain and in immune tissues, and the expression of *WNT3* and *MAP3* in immune tissues was significantly associated with ASD and lymphocyte and neutrophil count (p= 9.67x10-7- 1.22x10-14). We did not identify genes expressed in the brain and immune tissues significantly associated with ASD and immune phenotypes at the other shared loci.

### Enrichment of fetal mQTLs in shared loci

We registered a complete overlap (100%) of fetal mQTLs with ASD-related SNPs specifically falling with the locus (chr17:43-44Mb), where we registered a correlation between ASD and (respectively) AID, eosinophil and lymphocyte count (figure S1). At this locus, fetal brain mQTLs were associated with (cis) methylation at CpG islands at eight unique DNA locations corresponding to the *LRRC37A* and *MAPT* genes (Table S5). We did not identify any significant overlap between ASD-related SNPs and fetal mQTLs at the other shared loci (p > 0.05).

### Immune-based polygenic scores association with the autistic-like traits

PGS analyses indicated specific associations between genetic liability to immune-related phenotypes and autistic-like traits in a population-based sample (Table S6). The strongest association was reported between rigidity and PGS for SLE (best Pt=0.006; p_FDR_ = 0.03) (Figure 3; Table S7). Rigidity was also associated with PGSs for RA (Pt = 0.12; p_FDR_ =0.03) and ALG (Pt=0.052; p_FDR_ =0.03). In addition, we detected associations between the total autistic score and PGSs for ALG (Pt= 0.2; p_FDR_ =0.03), Lymph (Pt = 0.01; p_FDR_ = 0.03) and SLE (Pt= 0.0001; q=0.03) (Figure S2). Last, there was an association between childhood behaviour and PGS for LYMPH (Pt=0.0004; p_FDR_ =0.03; Figure 3). The association between immune-based PGS and the other tested autistic-like traits did not survive FDR-correction (p_FDR_ > 0.05). Also, sex-stratified associations between immune-based PGSs and autistic-like traits were not significant after FDR-correction (p_FDR_ > 0.05) (see Table S7; Figure S3-S5).

**Figure 3.**
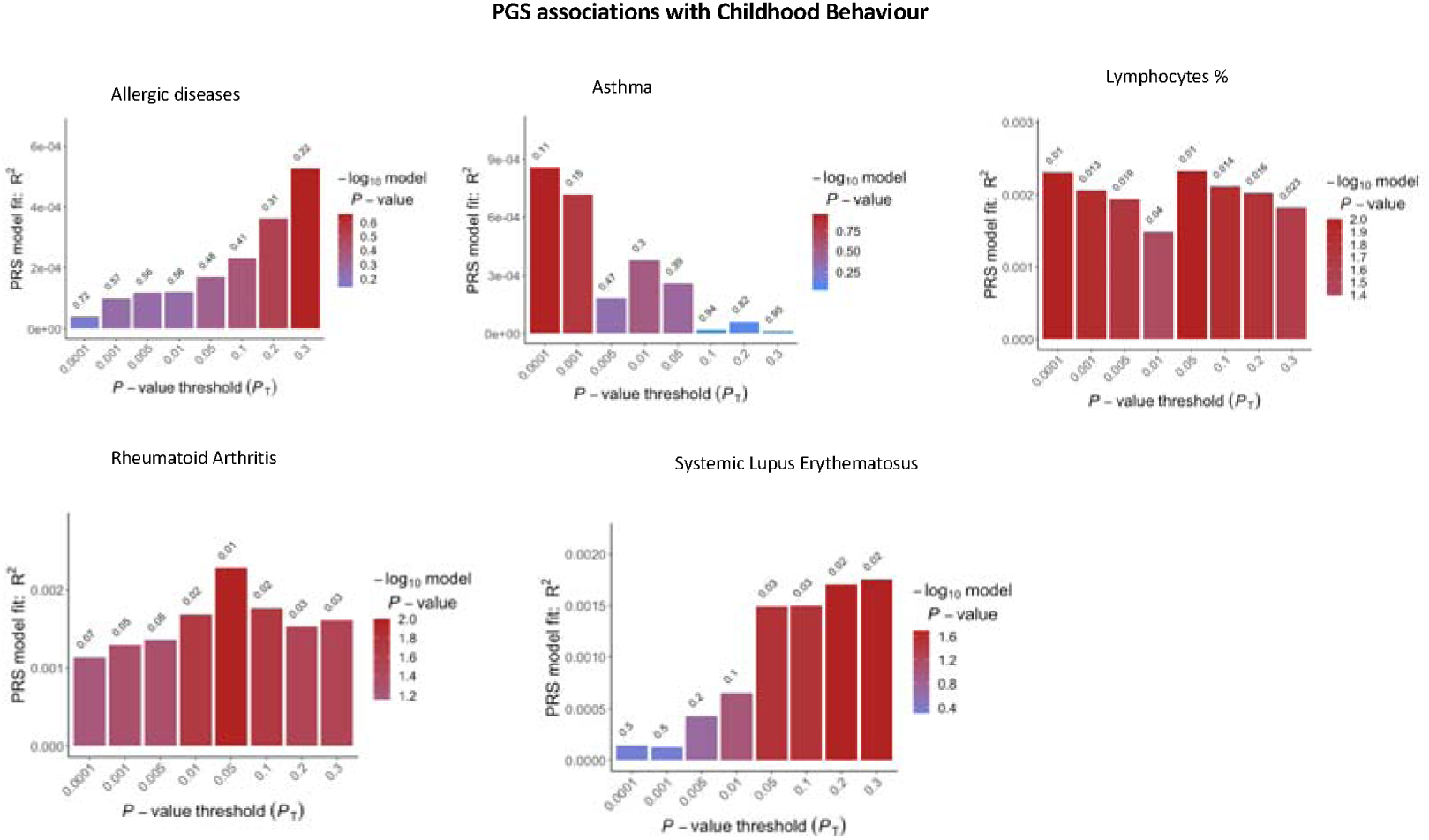
Bar plot results indicating the variance that polygenic scores for immune phenotypes associated with ASD explain in ‘childhood behaviour’.

## Discussion

In this study, we demonstrated that several autoimmune and atopic diseases share genetic liability with ASD. The genetic relationship between these immune phenotypes and ASD is complex, and its direction varies according to the specific immune phenotype considered. To further explore this genetic relationship, we investigated local genetic correlation and identified specific shared genomic loci. Some of these loci demonstrated an enrichment for common variants regulating gene expression in both immune tissues and brain; and which are involved in methylation during neurodevelopment. Furthermore, our results indicate that immunogenetic associations exist between specific autistic-like dimensions in general population.

Considering ASD as a clinical category, we reported a positive global genetic correlation between ASD and diseases associated with increased sensitivity to common allergens (i.e., allergies and asthma). These genetic correlations are in line with reports on the high prevalence of various allergic conditions in ASD (Miyazaki et al., 2015) and are consistent with prior findings of dysregulated expression of histamine signalling genes – key modulators of allergic reaction – in post-mortem ASD brains (Wright et al., 2017). In addition, we detected a negative global genetic correlation between ASD and lymphocyte count, which suggests that genetic factors associated with higher likelihood for ASD are also link to lower levels of peripheral lymphocytes, and *vice versa*. These findings, therefore, suggest the possibility of faulty adaptive, lymphocyte-mediated immune response. Notably, dysregulations in lymphocyte levels, and especially T cells, have been documented in the peripheral blood of autistic individuals (Ashwood et al., 2011). We also detected a negative genetic correlation between autoimmune conditions (RA and SLE) and ASD, suggesting that variants associated with increased likelihood ASD may be associated with resilience towards autoimmune diseases, and *vice versa*. These findings differ from epidemiological reports of a high rate of autoimmune conditions in the relatives of autistic individuals (Atladóttir et al., 2009). The evidence of both positive and negative genetic association suggest that ASD is linked to dysregulation in very specific immunogenetic mechanisms. To understand which of these mechanisms may be important, it is crucial to consider the aetiology of the immune phenotypes considered here. For example, prior studies suggest that allergic responses and autoimmunity may be ascribed both an imbalance between different classes of T helper (Th) lymphocytes, like Th1/Th17 and Th2 cells (Ashley et al., 2017; Bolon, 2012; Chaplin, 2010; Wahren-Herlenius & Dörner, 2013). However, while allergic responses have been associated with an increased Th2 cell activity as compared to Th1 cells, autoimmunity has been linked to a predominant Th1 response (Ashley et al., 2017; Bolon, 2012; Chaplin, 2010; Ramos et al., 2015). In this context, our findings suggest that deeper interrogation of whether ASD is linked to genetic factors regulating the Th1/Th2 homeostasis are warranted.

The negative genetic correlation findings reported here should be interpreted in the light of some methodological challenges. For example, the global LDSC based genetic correlation analyses exclude common HLA polymorphisms – due to the complex LD structure of the HLA region. This region is, however, central to the aetiology of most autoimmune conditions (B. Liu et al., 2021; Wahren-Herlenius & Dörner, 2013) and may further contribute to the co-occurrence of those conditions in ASD. Moreover, global genetic correlations analyses fail to detect scenarios in which the genetic correlation between two phenotypes varies (or has opposite directions) across different genomic regions, being masked when summed on a global scale (Werme et al., 2022). To overcome these limitations, we also assessed genetic correlation at the level of specific loci – including loci within the HLA region. The results of these analyses supported a role of HLA-specific SNPs encompassing the chr6:29Mb locus in the relationship between ASD and CRP. Genetic variants at this locus map to, and regulate, the expression of the *HLA-G* gene, which is known to intervene in the maternal-fetal interface and has been implicated in several neurodevelopmental conditions, including ASD (Guerini et al., 2019). When we examined HLA-loci in the relationship between ASD and autoimmunity, local genetic correlations at these loci did not survive multiple comparison correction, suggesting that other factors may drive the association between ASD and autoimmune diseases. A useful next step would be further studies that, for example, rely on *ad-hoc* imputation of HLA loci, to elucidate the influence of HLA-related SNPs on ASD.

Notably, the local genetic correlation approach also led to the identification of loci that are shared (pleiotropic) between ASD and multiple immune phenotypes. One of the loci with higher pleiotropy spans the chr17q21.31 region. This region on chromosome 17 includes an inversion polymorphism, which is common in the European population and that has been previously implicated in ASD and brain morphology (Adams et al., 2016; Ikram et al., 2012; Pain et al., 2019). Our analyses also indicated that variants at this locus influence expression in the brain and tissues of the immune system, suggesting a role of this genomic region in potential neuro-immune alterations. Last, we demonstrated that ASD-related variants in these regions act as mQTLs in the fetal brain, suggesting that these genetic factors may be important in the prenatal period and potentially interact with prenatal environmental challenges, including MIA and its cascade effects on brain development.

Another explanation for the complex pattern of correlations observed between ASD and immune phenotypes may be ascribed to the phenotypic heterogeneity of ASD. ASD is defined by different combinations of cognitive and behavioural symptoms (Georgiades et al., 2013). Prior work also suggested that these symptoms may be genetically distinct (Arenella et al., 2022; Warrier et al., 2019). Therefore, immunogenetic mechanisms may influence specific symptom domains, and these specific genetic effects may be diluted or transformed when adopting categorical definitions of ASD (Warrier et al., 2019). To address this point, we investigated if immunogenetic factors were associated with specific autistic dimensions or traits in the general population. We adopted a PGS approach, which considers the additive effect of common genetic variants across the genome, including the HLA region (S. W. Choi & O’Reilly, 2019). Our results demonstrate an association between immune-related genetic variations and rigidity and childhood behaviour. This is consistent with our prior work demonstrating an enrichment of SNPs associated with autistic-like traits, including rigidity and attention to detail, in eQTLs influencing the expression of immunogenetic pathways in human brain cortex (Arenella et al., 2022). Taken together our findings suggest that immunogenetic factors may particularly influence rigid cognitive/behavioural aspects of the autistic phenotype.

Last, potential confounder in the genetic relationship between ASD and autoimmune diseases is sex. This is because sex hormones differentially modulate the immune responses (Roved et al., 2017). In particular, testosterone is known to act as an immunosuppressant, whereas oestrogens have immunoregulatory properties (leading to autoimmunity in extreme cases) (Roved et al., 2017). As a result of these modulatory effects, immune diseases differentially affect the two sexes and are more prevalent in women (with an approximate female to male ratio of 10:1 (Wahren-Herlenius & Dörner, 2013). This sex-specific regulation suggests that immunogenetic factors may also have different effects on autistic phenotypes across sexes. One obstacle to the evaluation of the possibility of any inter-sex variability is that women are largely under-represented in the ASD clinical population due to higher prevalence of diagnosed ASD in men as compared to women (Halladay et al., 2015), but also the ’masking’ of ASD symptoms in females (Lockwood Estrin et al., 2020). However, the low female sample size in sex stratified GWAS of ASD did not allow us to test the relationship between ASD and immune phenotypes across sexes (Martin et al., 2021). To address this issue, we examined the association between immune-related genetic factors and autistic-like traits separately in women and men in a general population sample which had a balanced representation of both sexes. Our results from PGS analyses did not show significant sex-stratified associations. Larger female samples sizes in both general population and clinical ASD populations are required to investigate this question further.

Our work has both strengths and limitations. We explored the genetic relationship between ASD and the immune system, by leveraging the largest GWAS summary statistics for immune phenotypes being linked to different immunopathology (i.e., autoimmunity, atopy, and inflammation). In this regard, we used both categorical and dimensional approaches, as well as sex-stratified analyses, to disentangle the complex relationship between the immune system and autistic phenotypes. Another strength is the use of state-of-the-art genomic techniques to estimate global, and local genetic sharing between immune and autistic phenotypes. In contrast, one limitation of our study is the correlational/observational nature of our approach. Therefore, we cannot infer any causal role of immunogenetic factors in ASD. Moreover, we limited our analyses to immune phenotypes for which we could exploit well-powered GWAS data (i.e., based on sample size and h^2^_SNP_), and therefore we could not investigate other likely relevant immune phenotypes, like cytokine markers (including both Th1 and Th2- related cytokines) that may have provided further insights on ASD-linked immune mechanisms (Nath et al., 2019). In addition, our PGS-based findings – albeit reaching significance -demonstrated that immune-based PGSs only account for a relatively small proportion of phenotypic variance in the autistic-like traits, in line with other studies adopting the same methods (Den Braber et al., 2016). Furthermore, our study was restricted to European populations and therefore our findings cannot be generalised to other ethnicities.

## Conclusions

Our study demonstrates that genetic factors involved in autoimmunity and allergic responses may be important to ASD. However, while allergy-related genetic factors are associated with increased likelihood of having ASD, autoimmunity-related genetic factors link to reduced ASD likelihood. By leveraging different methods, we gain insights on i) genomic loci – and the genes within those - that register an association between ASD and immunity, and ii) specific autistic features, that – in the general population – associate with these immunogenetic factors. Overall, we demonstrated that immunogenetic factors, linked to ASD, may have a regulatory function in both the mature and the developing brain; and that these immunogenetic factors are specifically linked to autistic-like traits ‘rigidity’ and ‘childhood behaviour’.

## Data Availability

All data produced in the present work are contained in the manuscript.

https://pgc.unc.edu/for-the-public/information-on-disorders/autism-spectrum-disorder/

## References

1. Adams, H. H. H., Hibar, D. P., Chouraki, V., Stein, J. L., Nyquist, P. A., Rentería, M. E., Trompet, S., Arias-Vasquez, A., Seshadri, S., Desrivières, S., Beecham, A. H., Jahanshad, N., Wittfeld, K., van der Lee, S. J., Abramovic, L., Alhusaini, S., Amin, N., Andersson, M., Arfanakis, K., … Thompson, P. M. (2016). Novel genetic loci underlying human intracranial volume identified through genome-wide association. Nature Neuroscience, 19(12), 1569–1582. 10.1038/nn.4398

2. American Psychiatric Association. (2013). Diagnostic and Statistical Manual of Mental Disorders, 5.

3. Angum, F., Khan, T., Kaler, J., Siddiqui, L., & Hussain, A. (2020). The Prevalence of Autoimmune Disorders in Women: A Narrative Review. Cureus, 12(5). 10.7759/cureus.8094

4. Arenella, M., Cadby, G., de Witte, W., Jones, R. M., Whitehouse, A. J., Moses, E. K., Fornito, A., Bellgrove, M. A., Hawi, Z., Johnson, B., Tiego, J., Buitelaar, J. K., Kiemeney, L. A., Poelmans, G., & Bralten, J. (2022). Potential role for immune-related genes in autism spectrum disorders: Evidence from genome-wide association meta-analysis of autistic traits. AutismlJ: The International Journal of Research and Practice, 26(2), 361–372. 10.1177/13623613211019547

5. Ashley, S. E., Tan, H. LJT. T., Peters, R., Allen, K. J., Vuillermin, P., Dharmage, S. C., Tang, M. L. K., Koplin, J., Lowe, A., Ponsonby, A. LJL., Molloy, J., Matheson, M. C., Saffery, R., Ellis, J. A., & Martino, D. (2017). Genetic variation at the Th2 immune gene *<SCP>IL</SCP> 13* is associated with IgELJmediated paediatric food allergy. Clinical & Experimental Allergy, 47(8), 1032–1037. 10.1111/cea.12942

6. Ashwood, P., & van de Water, J. (2004). A Review of Autism and the Immune Response. Clinical and Developmental Immunology, 11(2), 165–174. 10.1080/10446670410001722096

7. Atladóttir, H. Ó., Pedersen, M. G., Thorsen, P., Mortensen, P. B., Deleuran, B., Eaton, W. W., & Parner, E. T. (2009). Association of family history of autoimmune diseases and autism spectrum disorders. Pediatrics, 124(2), 687–694. 10.1542/peds.2008-2445

8. Bennabi, M., Gaman, A., Delorme, R., Boukouaci, W., Manier, C., Scheid, I., Si Mohammed, N., Bengoufa, D., Charron, D., Krishnamoorthy, R., Leboyer, M., & Tamouza, R. (2018). HLA-class II haplotypes and Autism Spectrum Disorders. Scientific Reports, 8(1), 1–8. 10.1038/s41598-018-25974-9

9. Bentham, J., Morris, D. L., Cunninghame Graham, D. S., Pinder, C. L., Tombleson, P., Behrens, T. W., Martín, J., Fairfax, B. P., Knight, J. C., Chen, L., Replogle, J., Syvänen, A.-C., Rönnblom, L., Graham, R. R., Wither, J. E., Rioux, J. D., Alarcón-Riquelme, M. E., & Vyse, T. J. (2015). Genetic association analyses implicate aberrant regulation of innate and adaptive immunity genes in the pathogenesis of systemic lupus erythematosus Europe PMC Funders Group. Nat Genet, 47(12), 1457–1464. 10.1038/ng.3434

10. Bolon, B. (2012). Cellular and Molecular Mechanisms of Autoimmune Disease. Toxicologic Pathology, 40(2), 216–229. 10.1177/0192623311428481

11. Boulanger-Bertolus, J., Pancaro, C., & Mashour, G. A. (2018). Increasing role of maternal immune activation in neurodevelopmental disorders. Frontiers in Behavioral Neuroscience, 12(October), 1–6. 10.3389/fnbeh.2018.00230

12. Bralten, J., van Hulzen, K. J., Martens, M. B., Galesloot, T. E., Arias Vasquez, A., Kiemeney, L. A., Buitelaar, J. K., Muntjewerff, J. W., Franke, B., & Poelmans, G. (2018). Autism spectrum disorders and autistic traits share genetics and biology. Molecular Psychiatry, 23(5), 1205–1212. 10.1038/mp.2017.98

13. Bulik-Sullivan, B., Loh, P. R., Finucane, H. K., Ripke, S., Yang, J., Patterson, N., Daly, M. J., Price, A. L., Neale, B. M., Corvin, A., Walters, J. T. R., Farh, K. H., Holmans, P. A., Lee, P., Collier, D. A., Huang, H., Pers, T. H., Agartz, I., Agerbo, E., … O’Donovan, M. C. (2015). LD score regression distinguishes confounding from polygenicity in genome-wide association studies. Nature Genetics, 47(3), 291–295. 10.1038/ng.3211

14. Chaplin, D. D. (2010). Overview of the immune response. Journal of Allergy and Clinical Immunology, 125(2), S3–S23. 10.1016/j.jaci.2009.12.980

15. Chiarotti, F., & Venerosi, A. (2020). Epidemiology of autism spectrum disorders: A review of worldwide prevalence estimates since 2014. Brain Sciences, 10(5). 10.3390/brainsci10050274

16. Choi, G. B., Yim, Y. S., Wong, H., Kim, S., Kim, H., Kim, S. v., Hoeffer, C. A., Littman, D. R., & Huh, J. R. (2016). The maternal interleukin-17a pathway in mice promotes autism-like phenotypes in offspring. Science, 351(6276), 933–939. 10.1126/science.aad0314

17. Choi, S. W., & O’Reilly, P. F. (2019). PRSice-2: Polygenic Risk Score software for biobank-scale data. GigaScience, 8(7), 1–6. 10.1093/gigascience/giz082

18. Den Braber, A., Zilhão, N. R., Fedko, I. O., Hottenga, J. J., Pool, R., Smit, D. J. A., Cath, D. C., & Boomsma, D. I. (2016). Obsessive–compulsive symptoms in a large population-based twin-family sample are predicted by clinically based polygenic scores and by genome-wide SNPs. Translational Psychiatry, 6(2), 1–7. 10.1038/tp.2015.223

19. Dubois, P. C. A., Trynka, G., Franke, L., Hunt, K. A., Romanos, J., Curtotti, A., Zhernakova, A., Heap, G. A. R., Ádány, R., Aromaa, A., Bardella, M. T., van den Berg, L. H., Bockett, N. A., de la Concha, E. G., Dema, B., Fehrmann, R. S. N., Fernández-Arquero, M., Fiatal, S., Grandone, E., … van Heel, D. A. (2010). Multiple common variants for celiac disease influencing immune gene expression. Nature Genetics, 42(4), 295–302. 10.1038/ng.543

20. Ecker, C., Pretzsch, C. M., Bletsch, A., Mann, C., Schaefer, T., Ambrosino, S., Tillmann, J., Yousaf, A., Chiocchetti, A., Lombardo, M. v., Warrier, V., Bast, N., Moessnang, C., Baumeister, S., Dell’Acqua, F., Floris, D. L., Zabihi, M., Marquand, A., Cliquet, F., … Murphy, D. G. M. (2022). Interindividual Differences in Cortical Thickness and Their Genomic Underpinnings in Autism Spectrum Disorder. American Journal of Psychiatry, 179(3), 242–254. 10.1176/appi.ajp.2021.20050630

21. Edmiston, E., Ashwood, P., & van de Water, J. (2018). AUTOIMMUNITY, AUTOANTIBODIES, AND AUTISM SPECTRUM DISORDERS (ASD). Biological Psychiatry, 81(5), 383–390. 10.1016/j.biopsych.2016.08.031.AUTOIMMUNITY

22. Estes, M. L., & McAllister, A. K. (2016). Maternal immune activation: Implications for neuropsychiatric disorders. Science, 353(6301), 772–777. 10.1126/science.aag3194

23. Forgetta, V., Manousaki, D., Istomine, R., Ross, S., Tessier, M.-C., Marchand, L., Li, M., Qu, H.-Q., Bradfield, J. P., Grant, S. F. A., Hakonarson, H., DCCT/EDIC Research Group, Paterson, A. D., Piccirillo, C., Polychronakos, C., & Richards, J. B. (2020). Rare Genetic Variants of Large Effect Influence Risk of Type 1 Diabetes. Diabetes, 69(4), 784–795. 10.2337/db19-0831

24. Galesloot, T. E., Vermeulen, S. H., Swinkels, D. W., de Vegt, F., Franke, B., den Heijer, M., de Graaf, J., Verbeek, A. L. M., & Kiemeney, L. A. L. M. (2017). Cohort Profile: The Nijmegen Biomedical Study (NBS). International Journal of Epidemiology, 46(4), 1099–1100j. 10.1093/ije/dyw268

25. Gandal, M. J., Zhang, P., Hadjimichael, E., Walker, R. L., Chen, C., Liu, S., Won, H., van Bakel, H., Varghese, M., Wang, Y., Shieh, A. W., Haney, J., Parhami, S., Belmont, J., Kim, M., Losada, P. M., Khan, Z., Mleczko, J., Xia, Y., … Geschwind, D. H. (2018). Transcriptome-wide isoform-level dysregulation in ASD, schizophrenia, and bipolar disorder. Science, 362(6420). 10.1126/science.aat8127

26. Georgiades, S., Szatmari, P., Boyle, M., Hanna, S., Duku, E., Zwaigenbaum, L., Bryson, S., Fombonne, E., Volden, J., Mirenda, P., Smith, I., Roberts, W., Vaillancourt, T., Waddell, C., Bennett, T., & Thompson, A. (2013). Investigating phenotypic heterogeneity in children with autism spectrum disorder: A factor mixture modeling approach. Journal of Child Psychology and Psychiatry and Allied Disciplines, 54(2), 206–215. 10.1111/j.1469-7610.2012.02588.x

27. Gerring, Z. F., Mina-Vargas, A., Gamazon, E. R., & Derks, E. M. (2021). E-MAGMA: an eQTL-informed method to identify risk genes using genome-wide association study summary statistics. Bioinformatics, 37(16), 2245–2249. 10.1093/bioinformatics/btab115

28. Grove, J., Ripke, S., Als, T. D., Mattheisen, M., Walters, R. K., Won, H., Pallesen, J., Agerbo, E., Andreassen, O. A., Anney, R., Awashti, S., Belliveau, R., Bettella, F., Buxbaum, J. D., Bybjerg-Grauholm, J., Bækvad-Hansen, M., Cerrato, F., Chambert, K., Christensen, J. H., … Børglum, A. D. (2019). Identification of common genetic risk variants for autism spectrum disorder. Nature Genetics, 51(3), 431–444. 10.1038/s41588-019-0344-8

29. Halladay, A. K., Bishop, S., Constantino, J. N., Daniels, A. M., Koenig, K., Palmer, K., Messinger, D., Pelphrey, K., Sanders, S. J., Singer, A. T., Taylor, J. L., & Szatmari, P. (2015). Sex and gender differences in autism spectrum disorder: summarizing evidence gaps and identifying emerging areas of priority. Molecular Autism, 6(1), 36. 10.1186/s13229-015-0019-y

30. Han, X., Ong, J.-S., An, J., Hewitt, A. W., Gharahkhani, P., & MacGregor, S. (2020). Using Mendelian randomization to evaluate the causal relationship between serum C-reactive protein levels and age-related macular degeneration. European Journal of Epidemiology, 35(2), 139–146. 10.1007/s10654-019-00598-z

31. Han, Y., Jia, Q., Jahani, P. S., Hurrell, B. P., Pan, C., Huang, P., Gukasyan, J., Woodward, N. C., Eskin, E., Gilliland, F. D., Akbari, O., Hartiala, J. A., & Allayee, H. (2020). Genome-wide analysis highlights contribution of immune system pathways to the genetic architecture of asthma. Nature Communications, 11(1), 1776. 10.1038/s41467-020-15649-3

32. Hannon, E., Spiers, H., Viana, J., Pidsley, R., Burrage, J., Murphy, T. M., Troakes, C., Turecki, G., O’Donovan, M. C., Schalkwyk, L. C., Bray, N. J., & Mill, J. (2016). Methylation QTLs in the developing brain and their enrichment in schizophrenia risk loci. Nature Neuroscience, 19(1), 48–54. 10.1038/nn.4182

33. Ikram, M. A., Fornage, M., Smith, A. v, Seshadri, S., Schmidt, R., Debette, S., Vrooman, H. A., Sigurdsson, S., Ropele, S., Taal, H. R., Mook-Kanamori, D. O., Coker, L. H., Longstreth, W. T., Niessen, W. J., DeStefano, A. L., Beiser, A., Zijdenbos, A. P., Struchalin, M., Jack, C. R., … Cohorts for Heart and Aging Research in Genomic Epidemiology Consortium. (2012). Common variants at 6q22 and 17q21 are associated with intracranial volume. Nature Genetics, 44(5), 539–544. 10.1038/ng.2245

34. Liu, B., Shao, Y., & Fu, R. (2021). Current research status of HLA in immuneLJrelated diseases. *Immunity*, Inflammation and Disease, 9(2), 340–350. 10.1002/iid3.416

35. Liu, K., Huang, Y., Zhu, Y., Zhao, Y., & Kong, X. (2023). The role of maternal immune activation in immunological and neurological pathogenesis of autism. Journal of Neurorestoratology, 11(1), 100030. 10.1016/j.jnrt.2022.100030

36. Lockwood Estrin, G., Milner, V., Spain, D., Happé, F., & Colvert, E. (2020). Barriers to Autism Spectrum Disorder Diagnosis for Young Women and Girls: a Systematic Review. In Review Journal of Autism and Developmental Disorders. Springer. 10.1007/s40489-020-00225-8

37. Lombardo, M. v., Moon, H. M., Su, J., Palmer, T. D., Courchesne, E., & Pramparo, T. (2018). Maternal immune activation dysregulation of the fetal brain transcriptome and relevance to the pathophysiology of autism spectrum disorder. Molecular Psychiatry, 23(4), 1001–1013. 10.1038/mp.2017.15

38. Martin, J., Khramtsova, E. A., Goleva, S. B., Blokland, G. A. M., Traglia, M., Walters, R. K., Hübel, C., Coleman, J. R. I., Breen, G., Børglum, A. D., Demontis, D., Grove, J., Werge, T., Bralten, J., Bulik, C. M., Lee, P. H., Mathews, C. A., Peterson, R. E., Winham, S. J., … Stahl, E. (2021). Examining Sex-Differentiated Genetic Effects Across Neuropsychiatric and Behavioral Traits. Biological Psychiatry, 89(12), 1127–1137. 10.1016/j.biopsych.2020.12.024

39. Matoba, N., Liang, D., Sun, H., Aygün, N., McAfee, J. C., Davis, J. E., Raffield, L. M., Qian, H., Piven, J., Li, Y., Kosuri, S., Won, H., & Stein, J. L. (2020). Common genetic risk variants identified in the SPARK cohort support DDHD2 as a candidate risk gene for autism. Translational Psychiatry, 10(1). 10.1038/s41398-020-00953-9

40. McAllister, A. K. (2017). Immune contributions to cause and effect in autism spectrum disorder. Biological Psychiatry, 81(5), 380–382. 10.1111/mec.13536.Application

41. Miyazaki, C., Koyama, M., Ota, E., Swa, T., Amiya, R. M., Mlunde, L. B., Tachibana, Y., Yamamoto-Hanada, K., & Mori, R. (2015). Allergies in Children with Autism Spectrum Disorder: a Systematic Review and Meta-analysis. Review Journal of Autism and Developmental Disorders, 2(4), 374–401. 10.1007/s40489-015-0059-4

42. Mostafa, G. A., Shehab, A. A., & Al-Ayadhi, L. Y. (2013). The link between some alleles on human leukocyte antigen system and autism in children. Journal of Neuroimmunology, 255(1–2), 70–74. 10.1016/j.jneuroim.2012.10.002

43. Nath, A. P., Ritchie, S. C., Grinberg, N. F., Tang, H. H.-F., Huang, Q. Q., Teo, S. M., Ahola-Olli, A. v, Würtz, P., Havulinna, A. S., Santalahti, K., Pitkänen, N., Lehtimäki, T., Kähönen, M., Lyytikäinen, L.-P., Raitoharju, E., Seppälä, I., Sarin, A.-P., Ripatti, S., Palotie, A., … Inouye, M. (2019). Multivariate Genome-wide Association Analysis of a Cytokine Network Reveals Variants with Widespread Immune, Haematological, and Cardiometabolic Pleiotropy. American Journal of Human Genetics, 105(6), 1076–1090. 10.1016/j.ajhg.2019.10.001

44. Okada, Y., Wu, D., Trynka, G., Raj, T., Terao, C., Ikari, K., Kochi, Y., Ohmura, K., Suzuki, A., Yoshida, S., Graham, R. R., Manoharan, A., Ortmann, W., Bhangale, T., Denny, J. C., Carroll, R. J., Eyler, A. E., Greenberg, J. D., Kremer, J. M., … Plenge, R. M. (2014). Genetics of rheumatoid arthritis contributes to biology and drug discovery. Nature, 506(7488), 376–381. 10.1038/nature12873

45. Pain, O., Pocklington, A. J., Holmans, P. A., Bray, N. J., O’Brien, H. E., Hall, L. S., Pardiñas, A. F., O’Donovan, M. C., Owen, M. J., & Anney, R. (2019). Novel Insight Into the Etiology of Autism Spectrum Disorder Gained by Integrating Expression Data With Genome-wide Association Statistics. Biological Psychiatry, 86(4), 265–273. 10.1016/j.biopsych.2019.04.034

46. Purcell, S., Neale, B., Todd-Brown, K., Thomas, L., Ferreira, M. A. R., Bender, D., Maller, J., Sklar, P., de Bakker, P. I. W., Daly, M. J., & Sham, P. C. (2007). PLINK: A Tool Set for Whole-Genome Association and Population-Based Linkage Analyses. The American Journal of Human Genetics, 81(3), 559–575. 10.1086/519795

47. Ramos, P. S., Shedlock, A. M., & Langefeld, C. D. (2015). Genetics of autoimmune diseases: insights from population genetics. Journal of Human Genetics, 60(11), 657–664. 10.1038/jhg.2015.94

48. Rogge, N., & Janssen, J. (2019). The Economic Costs of Autism Spectrum Disorder: A Literature Review. Journal of Autism and Developmental Disorders, 49(7), 2873–2900. 10.1007/s10803-019-04014-z

49. Roved, J., Westerdahl, H., & Hasselquist, D. (2017). Sex differences in immune responses: Hormonal effects, antagonistic selection, and evolutionary consequences. Hormones and Behavior, 88, 95–105. 10.1016/j.yhbeh.2016.11.017

50. Saevarsdottir, S., Olafsdottir, T. A., Ivarsdottir, E. v., Halldorsson, G. H., Gunnarsdottir, K., Sigurdsson, A., Johannesson, A., Sigurdsson, J. K., Juliusdottir, T., Lund, S. H., Arnthorsson, A. O., Styrmisdottir, E. L., Gudmundsson, J., Grondal, G. M., Steinsson, K., Alfredsson, L., Askling, J., Benediktsson, R., Bjarnason, R., … Stefansson, K. (2020). FLT3 stop mutation increases FLT3 ligand level and risk of autoimmune thyroid disease. Nature, 584(7822), 619– 623. 10.1038/s41586-020-2436-0

51. Smith, S. E. P., Li, J., Garbett, K., Mirnics, K., & Patterson, P. H. (2007). Maternal Immune Activation Alters Fetal Brain Development through Interleukin-6. Journal of Neuroscience, 27(40), 10695–10702. 10.1523/JNEUROSCI.2178-07.2007

52. Torres, A. R., Sweeten, T. L., Johnson, R. C., Odell, D., Westover, J. B., Bray-Ward, P., Ward, D. C., Davies, C. J., Thomas, A. J., Croen, L. A., & Benson, M. (2016). Common genetic variants found in HLA and KIR immune genes in autism spectrum disorder. Frontiers in Neuroscience, 10(OCT), 1–13. 10.3389/fnins.2016.00463

53. Traglia, M., Croen, L. A., Jones, K. L., Heuer, L. S., Yolken, R., Kharrazi, M., DeLorenze, G. N., Ashwood, P., van de Water, J., & Weiss, L. A. (2018). Cross-genetic determination of maternal and neonatal immune mediators during pregnancy. Genome Medicine, 10(1), 1–17. 10.1186/s13073-018-0576-8

54. van Belle, G., Fisher, L. D., Heagerty, P. J., & Lumley, T. (2004). *Biostatistics*. John Wiley & Sons, Inc. 10.1002/0471602396

55. van Heijst, B., & Geurts, H. (2015). Quality of life in autism across the lifespan: a meta-analysis. Autism, 19(2), 158–167.

56. Vuckovic, D., Bao, E. L., Akbari, P., Lareau, C. A., Mousas, A., Jiang, T., Chen, M.-H., Raffield, L. M., Tardaguila, M., Huffman, J. E., Ritchie, S. C., Megy, K., Ponstingl, H., Penkett, C. J., Albers, P. K., Wigdor, E. M., Sakaue, S., Moscati, A., Manansala, R., … Soranzo, N. (2020). The Polygenic and Monogenic Basis of Blood Traits and Diseases. Cell, 182(5), 1214–1231.e11. 10.1016/j.cell.2020.08.008

57. Wahren-Herlenius, M., & Dörner, T. (2013). Immunopathogenic mechanisms of systemic autoimmune disease. The Lancet, 382(9894), 819–831. 10.1016/S0140-6736(13)60954-X

58. Warrier, V., Toro, R., Won, H., Leblond, C. S., Cliquet, F., Delorme, R., de Witte, W., Bralten, J., Chakrabarti, B., Børglum, A. D., Grove, J., Poelmans, G., Hinds, D. A., Bourgeron, T., & Baron-Cohen, S. (2019). Social and non-social autism symptoms and trait domains are genetically dissociable. Communications Biology, 2(1), 328. 10.1038/s42003-019-0558-4

59. Werme, J., van der Sluis, S., Posthuma, D., & de Leeuw, C. A. (2022). An integrated framework for local genetic correlation analysis. Nature Genetics, 54(3), 274–282. 10.1038/s41588-022-01017-y

60. Zerbo, O., Leong, A., Barcellos, L., Bernal, P., Fireman, B., & Croen, L. A. (2015). Immune mediated conditions in autism spectrum disorders. *Brain*, Behavior, and Immunity, 46, 232–236. 10.1016/j.bbi.2015.02.001

61. Zheng, J., Erzurumluoglu, A. M., Elsworth, B. L., Kemp, J. P., Howe, L., Haycock, P. C., Hemani, G., Tansey, K., Laurin, C., Pourcain, B. st., Warrington, N. M., Finucane, H. K., Price, A. L., Bulik-Sullivan, B. K., Anttila, V., Paternoster, L., Gaunt, T. R., Evans, D. M., & Neale, B. M. (2017). LD Hub: a centralized database and web interface to perform LD score regression that maximizes the potential of summary level GWAS data for SNP heritability and genetic correlation analysis. Bioinformatics, 33(2), 272–279. 10.1093/bioinformatics/btw613

62. Zhu, Z., Lee, P. H., Chaffin, M. D., Chung, W., Loh, P.-R., Lu, Q., Christiani, D. C., & Liang, L. (2018). A genome-wide cross-trait analysis from UK Biobank highlights the shared genetic architecture of asthma and allergic diseases. Nature Genetics, 50(6), 857–864. 10.1038/s41588-018-0121-0

