## Supplementary materials for "Genetic relationship between the immune system and autism spectrum disorder and traits"

### Supplementary material

Table S 1 Questions used in the Nijmegen Biomedical Study to measure autistic-like traits

**Questions used to measure four autistic-like traits**

**Attention to detail**

By looking at someone face, I find easy to work out what is he or she is thinking or feeling

I can quickly workout whether someone is fascinated by what I say

I tend to notice details that others do not ₐ

**Imagination**

I find making stories up easy

As a child, I enjoyed playing games involving pretending with other children

**Rigidity**

People tell me that I keep going on and on about the same thing ₐ

I often get so absorbed that I lose sight of other things ₐ

It upsets me if my daily routine is disturbed ₐ

I prefer to do things the same way over and over again ₐ

**Social skills**

I find it hard to make new friends ₐ

I enjoy social occasions as birthdays, receptions, etc.

I don’t know how to keep a conversation going ₐ

**Childhood behaviour**

As a child, I was a late talker, or I had other speech-related problems ₐ

As a child, I often retreated to my own world, or I rarely played with other children ₐ

As a child, I moved in a rigid way, or I tended to repeat certain movements ₐ

As a child, I often repeated the same words, or I made up new words ₐ

As a child, I often took statements and jokes literally ₐ

As a child, I frequently moved became upset by sudden and unexpected changes ₐ

Table S 2 Matrix showing the genetic inter-correlation between the different immune phenotypes

|  | **ast** | **alg** | **aid** | **cd** | **crp** | **eos** | **lymph** | **monoc** | **neutr** | **ra** | **sle** | **t1d** |
| --- | --- | --- | --- | --- | --- | --- | --- | --- | --- | --- | --- | --- |
| **ast** | 1,00 | 0,80 | 0,05 | 0,11 | 0,15 | 0,35 | -0,07 | -0,06 | -0,05 | 0,08 | 0,01 | 0,03 |
| **alg** | 0,80 | 1,00 | 0,07 | 0,32 | 0,01 | 0,33 | -0,05 | -0,03 | -0,07 | -0,01 | -0,07 | 0,06 |
| **aid** | 0,05 | 0,07 | 1,00 | 0,30 | 0,12 | 0,12 | -0,01 | 0,01 | -0,20 | 0,40 | 0,22 | 0,57 |
| **cd** | 0,11 | 0,32 | 0,30 | 1,00 | -0,07 | 0,09 | -0,09 | 0,01 | -0,15 | 0,18 | 0,16 | 0,20 |
| **crp** | 0,15 | 0,01 | 0,12 | -0,07 | 1,00 | 0,07 | -0,06 | -0,08 | -0,04 | 0,15 | 0,07 | -0,04 |
| **eos** | 0,35 | 0,33 | 0,12 | 0,09 | 0,07 | 1,00 | 0,01 | 0,01 | 0,02 | 0,09 | 0,05 | 0,22 |
| **lymph** | -0,07 | -0,05 | -0,01 | -0,09 | -0,06 | 0,01 | 1,00 | 0,01 | 0,97 | -0,01 | 0,01 | -0,03 |
| **monoc** | -0,06 | -0,03 | 0,01 | 0,01 | -0,08 | 0,01 | 0,01 | 1,00 | 0,03 | -0,02 | -0,02 | 0,04 |
| **neutr** | -0,05 | -0,07 | 0,20 | -0,15 | -0,04 | 0,02 | 0,97 | 0,03 | 1,00 | -0,08 | -0,04 | 0,05 |
| **ra** | 0,08 | -0,01 | 0,40 | 0,18 | 0,15 | 0,09 | -0,01 | -0,02 | -0,08 | 1,00 | 0,47 | 0,50 |
| **sle** | 0,01 | -0,07 | 0,22 | 0,16 | 0,07 | 0,05 | 0,01 | -0,02 | -0,04 | 0,47 | 1,00 | 0,24 |
| **t1d** | 0,03 | 0,06 | 0,57 | 0,20 | -0,04 | 0,22 | -0,03 | 0,04 | 0,05 | 0,50 | 0,24 | 1,00 |
| *Abbreviations : ast = asthma; alg = allergic disease; aid = autoimmune thyroid diseases; cd = celiac disease; crp = c-reactive protein; eos = eosinophil count; lymph = lymphocyte count; monoc = monocyte count; neutr = neutrophil count; ra = rheumatoid arthritis; sle = systemic lupus erythematosus; t1d = autoimmune type 1 diabetes.* | | | | | | | | | | | | |

Table S 3 Global genetic correlation results between immune phenotypes and ASD and number of shared loci

| Immune phenotypes | rg | se | p | q | N loci with significant h^2^_SNP_ |
| --- | --- | --- | --- | --- | --- |
| Allergy | 0.14 | 0.04 | 0.006 | 0.01 | 21 |
| Asthma | 0.08 | 0.03 | 0.01 | 0.02 | 17 |
| AID | 0.09 | 0.04 | 0.03 | 0.06 | 9 |
| CRP | 0.0008 | 0.02 | 0.97 | 0.97 | 37 |
| CD | 0.11 | 0.07 | 0.11 | 0.18 | 110 |
| Eos | 0.02 | 0.03 | 0.40 | 0.57 | 34 |
| Lymph | -0.06 | 0.02 | 0.005 | 0.01 | 21 |
| Monoc | 0.002 | 0.02 | 0.93 | 0.97 | 20 |
| Neutr | 0.02 | 0.03 | 0.55 | 0.66 | 78 |
| RA | -0.12 | 0.04 | 0.005 | 0.01 | 17 |
| SLE | -0.17 | 0.06 | 0.004 | 0.01 | 34 |
| T1D | 0.058 | 0.074 | 0.43 | 0.57 | 27 |
| *Abbreviations : AID = autoimmune thyroid diseases; HLA= human leukocyte antigen; CD = celiac disease; CRP = c-reactive protein; Eos = eosinophil count; Lymph = lymphocyte count; Monoc = monocyte count; Neutr = neutrophil count; RA = rheumatoid arthritis; SLE = systemic lupus* erythematosus; *T1D = autoimmune type 1 diabetes.; rg = genetic correlation; se = standard error; p = p-value; q = q-value;* h^2^_SNP_ *= SNP-based heritability* | | | | | |

Table S 4 Loci-specific genes expressed in the brain and immune system significantly associated with ASD and immune phenotypes

| Locus | e-Gene | Tissues | Trait-pair (p1-p2) | Z_p1 | P_p1 | Z_p2 | P_p2 |
| --- | --- | --- | --- | --- | --- | --- | --- |
| Locus chr 1 | *DDX9* | Transformed lymphocytes | ASD-Eos | 3.07 | 0.001 | 1.71 | 0.04 |
|  | *-* | spleen |  | 3.04 | 0.001 | 1.99 | 0.02 |
|  | *-* | Blood |  | 3.09 | 0.0009 | 2.12 | 0.01 |
| Locus chr 6 | *RNF39* | Brain cortex | ASD-CRP | 1.9 | 0.02 | 1.79 | 0.03 |
|  | *HLA-F* | Blood |  | 2.37 | 0.008 | 3.28 | 0.0005 |
|  | *HLA-F* | Transformed lymphocytes |  | 2.01 | 0.02 | 3.28 | 0.0005 |
|  | *HLA-A* |  |  |  |  |  |  |
|  | *ZFP57* | Transformed lymphocytes |  | 1.75 | 0.03 | e4.01 | 2.9x10-5 |
| Locus chr 12 | *YEATS4* | Brain cortex | ASD-Mon | 2.05 | 0.01 | 3.63 | 0.0001 |
|  |  | Blood |  | 3.17 | 0.0007 | 4.16 | 1.5x10-5 |
|  |  | Transformed lymphocytes |  | 2.47 | 0.006 | 3.59 | 0.0001 |
|  | *LYZ* | blood |  | 3.00 | 0.001 | 5.4 | 3.1x10-8 |
|  |  | spleen |  | 2.33 | 0.006 | 3.6 | 0.0001 |
|  | *MDM2* | blood |  | 1.17 | 0.01 | 3.78 | 7.8x10-5 |
| Locus chr 17 | *KANSL1* | blood | ASD-lymph/ntr | 4.86 | 5.7x10-7 | 7.8 | 2.8x10-15 |
|  | *ARL17A* | blood |  | 4.81 | 7.2x10-7 | 7.75 | 4.59x10-15 |
|  |  | spleen |  | 4.7 | 8.9x10-7 | 7.6 | 1.03x10-14 |
|  |  | Brain cortex |  | 4.83 | 6.73x10-7 | 7.85 | 2.05x10-15 |
|  | *LRRC37A* | blood |  | 4.86 | 5.7x10-7 | 7.89 | 1.4x10-15 |
|  |  | Spleen |  | 4.9 | 4.26x10-7 | 8.6 | 3.08x10-18 |
|  |  | Brain cortex |  | 4.8 | 6.2x10-7 | 7.9 | 1.21x10-15 |
|  | *LRRC37A2* | blood |  | 4.9 | 2.9x10-7 | 8.0 | 2.76x10-15 |
|  | *-* | Transformed lymphocytes |  | 4.76 | 9.67x10-7 | 7.73 | 5.4x10-14 |
|  |  | Spleen |  | 4.79 | 7.9x10-7 | 7.78 | 3.4x10-15 |
|  | *-* | Brain cortex |  | 4.81 | 7.4x10-7 | 7.97 | 7.836x10-16 |
|  | *WNT3* | Transformed lymphocytes |  | 4.56 | 2.5x10-7 | 7.6 | 1.22x10-14 |
|  |  | Spleen |  | 2.5 | 0.003 | 7.2 | 2.5x10-13 |
|  | *MAPT* | Spleen |  | 4.5 | 2.9x10-6 | 7.4 | 4.8x10-14 |
| *Abbreviations: chr = chromosome; Z = standardised effect size; p = pvalue; p1 = phenotype 1; p2 = phenotype 2; ASD = autism spectrum disorder; Eos = eosinophil count; Mon = monocyte count; CRP = c-reactive protein; Lymph = lymphocyte count; Neutr = neutrophil count;* | | | | | | | |

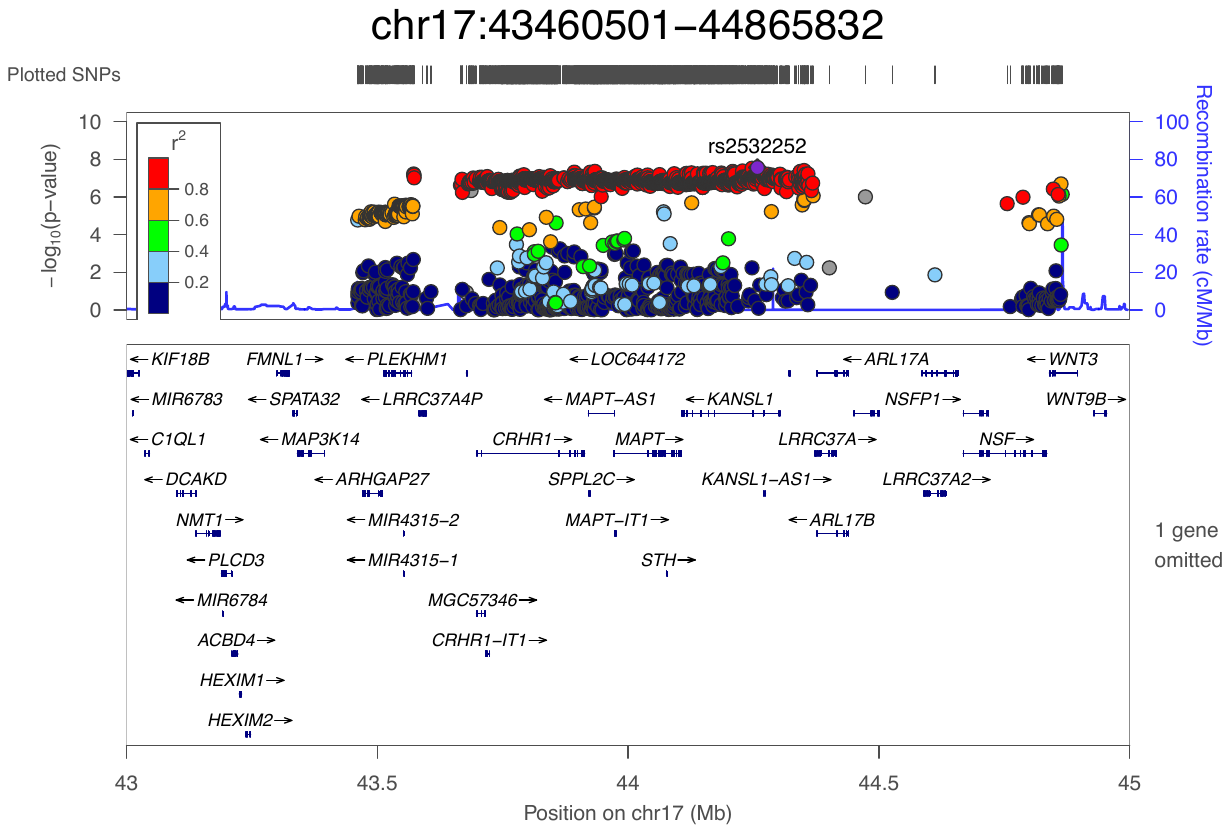

Figure S 1. Locus with rg signal between ASD and Autoimmune Thryoid diseases, Eosinophil count, Lymphocyte count. This locus registered a complete overlap between associated SNPs and significant fetal mQTLs.

Table S 5 Genomic locations of epigenetic modifications in the fetal brain associated overlapping with ASD-related SNPs at the locus chr17:43-44Mb.

| ***Site of DNA modification (chr:bp)*** | ***Gene*** | ***Gene location*** |
| --- | --- | --- |
| 17:43662623 | *LRRC37A* | TSS |
| 17:43662625 | *LRRC37A* | TSS |
| 17:43663208 | *LRRC37A* | TSS |
| 17:43663579 | *LRRC37A* | TSS |
| 17:43971911 | *MAPT* | TSS promoter |
| 17:43971919 | *MAPT* | TSS promoter |
| 17:43972573 | *MAPT* | Enhancer |
| 17:43973522 | *MAPT* | Enhancer  Low activity region |
| *Abbreviations: chr = chromosome; bp = basepair; TSS = transcription starting site;* | | |

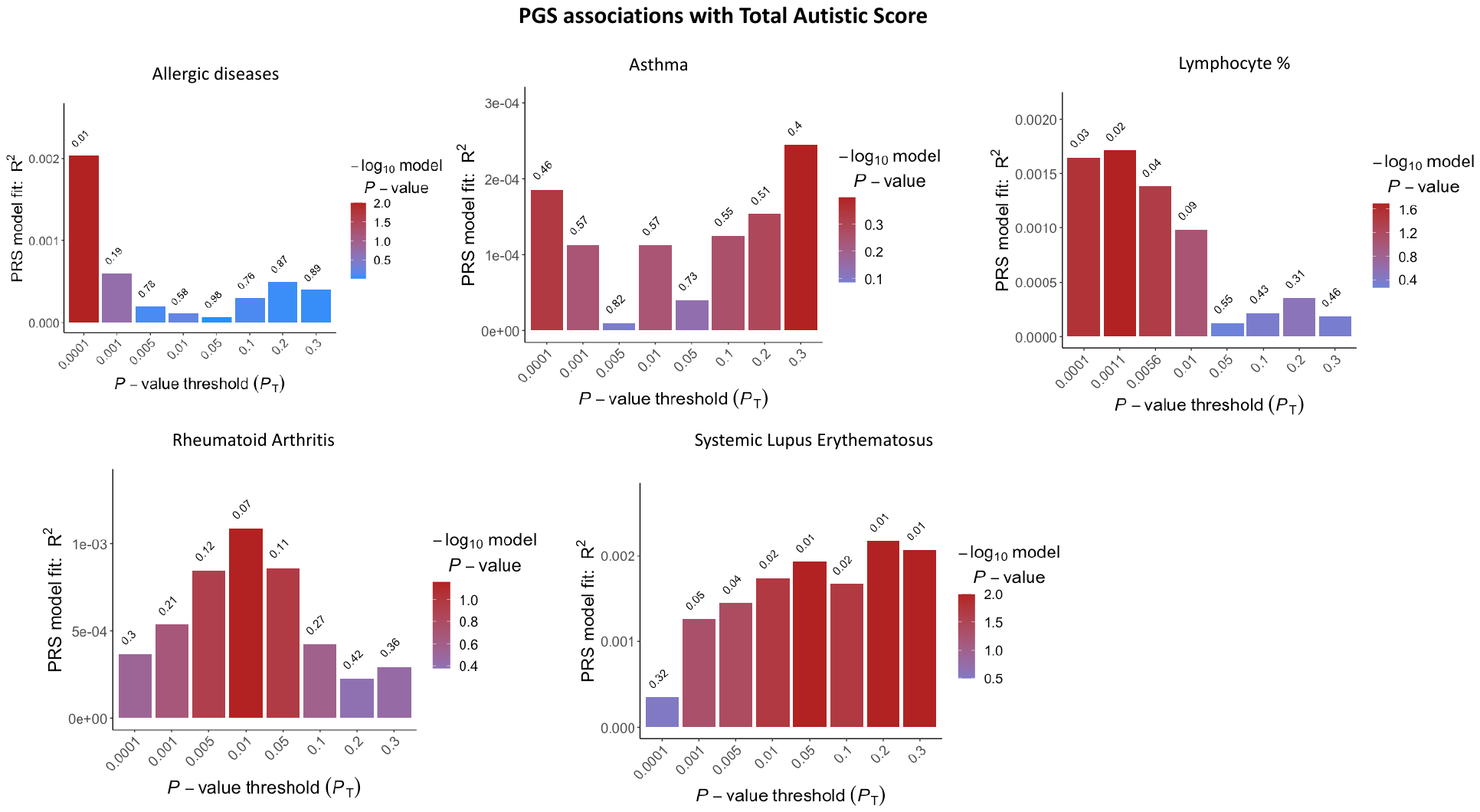

Figure S 2 Bar plot results indicating the variance that Polygenic scores for immune phenotypes associated with ASD explain in ‘total autistic score’.. The bars represent the aggregate sum of SNPs associated with each immune phenotype at different p-value threshold (pT) listed on the x-axis. The hight of the bar (y-axis) represents the degree of variance explained by each PT in ‘childhood behavior’. The colour of the bar defines the strength of the association and the significance by which rejecting the null hypothesis of no association between polygenic scores and ‘childhood behavior’. The p-value of association for each PT is reported on the top of each bar

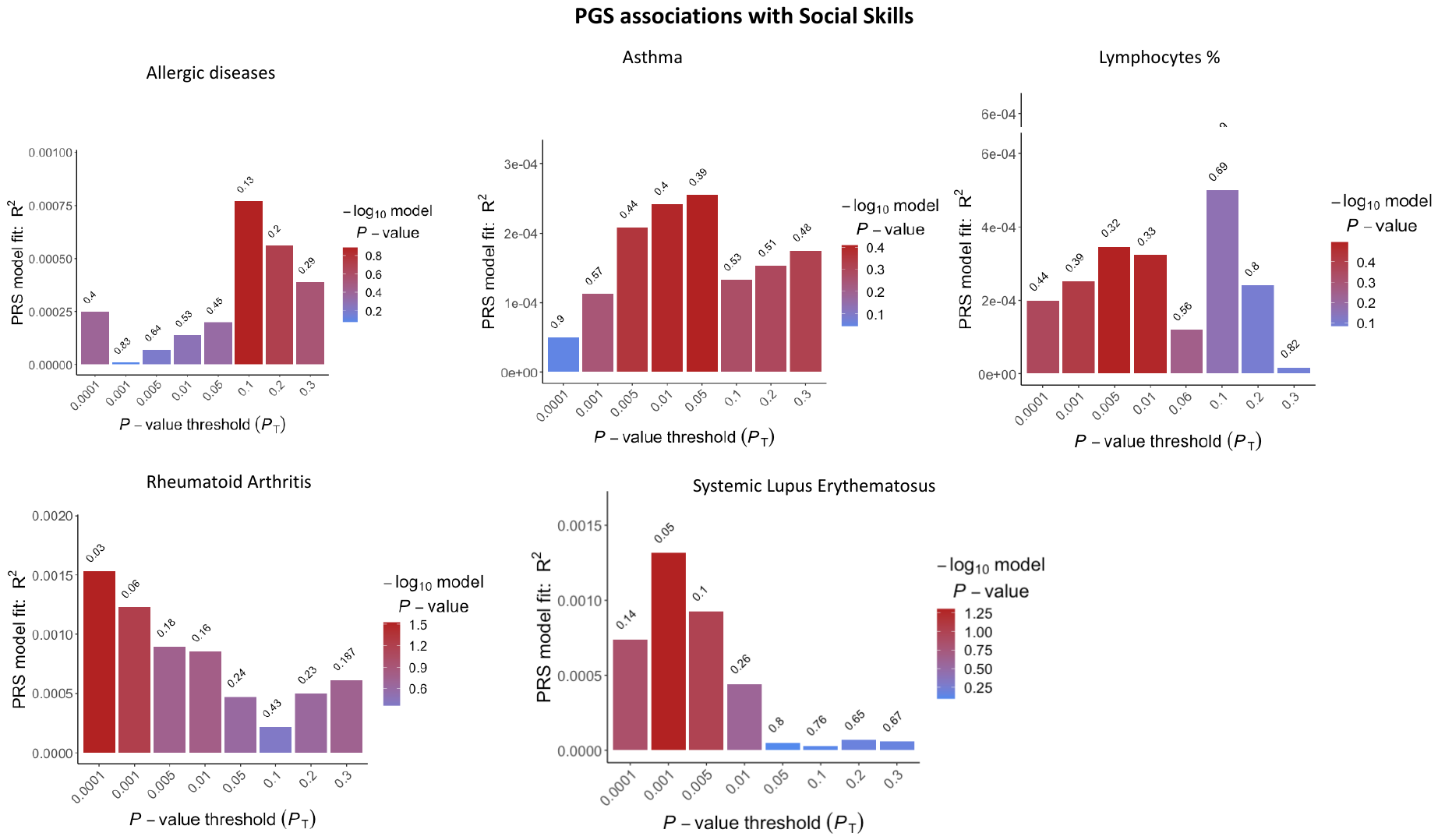

Figure S 3. Bar plot results indicating the variance that Polygenic scores for immune phenotypes associated with ASD explain in 'social skills'. The bars represent the aggregate sum of SNPs associated with each immune phenotype at different p-value threshold (pT) listed on the x-axis. The hight of the bar (y-axis) represents the degree of variance explained by each PT in ‘social skills’. The colour of the bar defines the strength of the association and the significance by which rejecting the null hypothesis of no association between polygenic scores and ‘social skills’. The p-value of association for each PT is reported on the top of each bar.

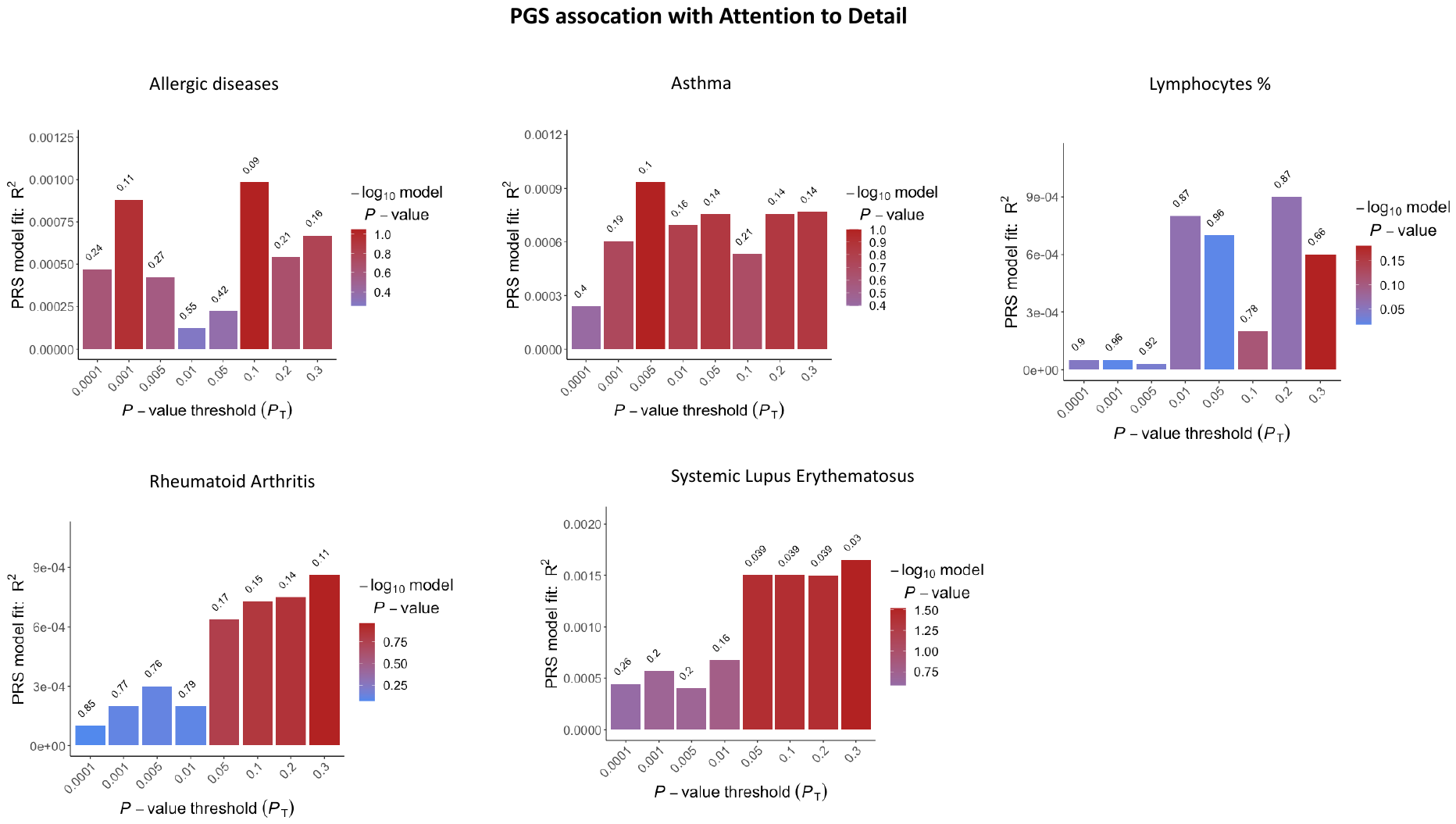

Figure S 4. Bar plot results indicating the variance that Polygenic scores for immune phenotypes associated with ASD explain in 'attention to detail'. The bars represent the aggregate sum of SNPs associated with each immune phenotype at different p-value threshold (pT) listed on the x-axis. The hight of the bar (y-axis) represents the degree of variance explained by each PT in ‘attention to detail’. The colour of the bar defines the strength of the association and the significance by which rejecting the null hypothesis of no association between polygenic scores and ‘attention to detail’. The p-value of association for each PT is reported on the top of each bar.

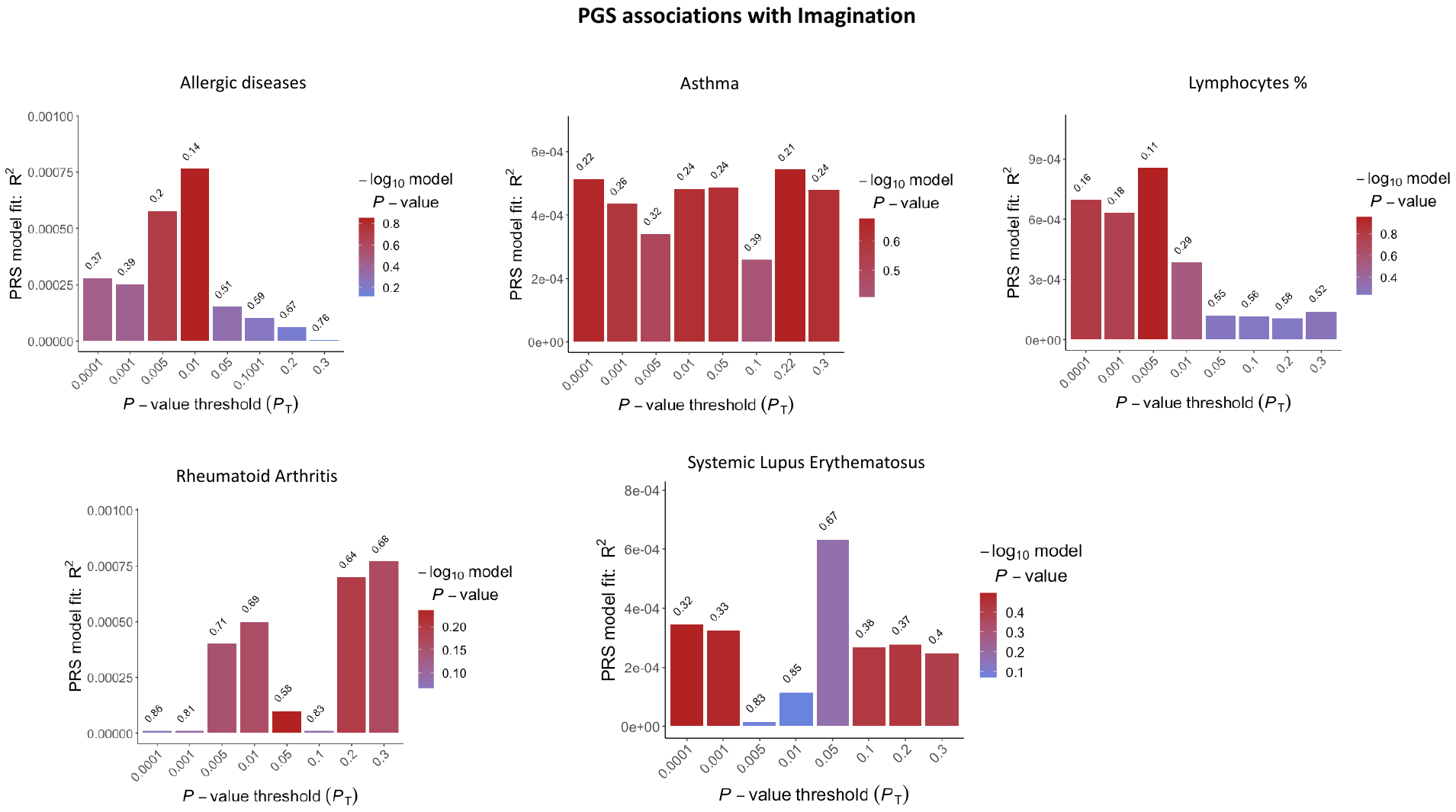

Figure S 5 Bar plot results indicating the variance that Polygenic scores for immune phenotypes associated with ASD explain in 'imagination'. The bars represent the aggregate sum of SNPs associated with each immune phenotype at different p-value threshold (pT) listed on the x-axis. The hight of the bar (y-axis) represents the degree of variance explained by each PT in ‘imagination’. The colour of the bar defines the strength of the association and the significance by which rejecting the null hypothesis of no association between polygenic scores and ‘imagination’. The p-value of association for each PT is reported on the top of each bar.

Table S 6 Results of regression analyses of polygenic score for immune phenotypes on each autistic-like traits

| Autistic-like traits |  |  |  |  |  |  |
| --- | --- | --- | --- | --- | --- | --- |
| Childhood behavior | Base GWAS | threshold | Full R2 | beta (se) | p-value | FDR-p value |
|  | ALG | 0.0001 | 0.0024 | -5.12(0.29) | 0.07 | 0.13 |
|  | AST | 1 | 0.0006 | 3.31(0.24) | 0.18 | 0.24 |
|  | **Lymph** | **0.0004** | **0.004** | **8.0(0.028)** | **0.005** | **0.03** |
|  | RA | 0.009 | 0.0015 | 3.9(0.03) | 0.37 | 0.39 |
|  | **SLE** | **1** | **0.0034** | **1.23(0.06)** | **0.01** | **0.03** |
| Rigidity |  |  |  |  |  |  |
|  | **ALG** | **0.13** | **0.0045** | **1.59(0.059)** | **0.007** | **0.03** |
|  | AST | 1 | 0.0030 | 4.16(0.23) | 0.07 | 0.13 |
|  | Lymph | 0.003 | 0.0023 | 5.2(0.49) | 0.28 | 0.32 |
|  | **RA** | **0.052** | **0.0043** | **2.5(0.09)** | **0.009** | **0.03** |
|  | **SLE** | **0.006** | **0.0051** | **1.1(0.03)** | **0.002** | **0.03** |
| Social skills |  |  |  |  |  |  |
|  | ALG |  |  |  |  |  |
|  | AST | 0.04 | 0.0026 | 7.56(0.71) | 0.2 | 0.26 |
|  | Lymph | 0.01 | 0.0026 | 1.3(0.11) | 0.21 | 0.27 |
|  | RA | 0.005 | 0.0037 | -2.5(0.15) | 0.11 | 0.16 |
|  | SLE | 0.001 | 0.0035 | -3.5(0.17) | 0.04 | 0.11 |
| Attention |  |  |  |  |  |  |
|  | ALG | 0.09 | 0.0012 | 5.93(0.37) | 0.07 | 0.13 |
|  | AST | 0.008 | 0.0013 | 2.9(0.16) | 0.07 | 0.13 |
|  | Lymph | 0.01 | 0.0003 | 3.4(0.61) | 0.57 | 0.58 |
|  | RA | 0.24 | 0.0010 | -2.5(0.15) | 0.11 | 0.16 |
|  | SLE | 1 | 0.0020 | -7.2(0.31) | 0.02 | 0.06 |
| Imagination |  |  |  |  |  |  |
|  | ALG | 0.01 | 0.0009 | 4.29(0.26) | 0.10 | 0.16 |
|  | AST | 0.0006 | 0.0013 | 1.4(0.10) | 0.17 | 0.24 |
|  | Lymph | 0.005 | 0.0017 | 1.1(0.07) | 0.11 | 0.16 |
|  | RA | 0.05 | 0.00010 | -6.5(0.51) | 0.58 | 0.58 |
|  | SLE | 0.17 | 0.0013 | -3.0(0.26) | 0.23 | 0.27 |
| Total |  |  |  |  |  |  |
|  | **ALG** | **0.00010** | **0.0035** | **-3.4(0.14)** | **0.01** | **0.03** |
|  | AST | 1 | 0.0019 | 1.43(0.13) | 0.30 | 0.33 |
|  | **Lymph** | **0.0004** | **0.0035** | **3.8(0.16)** | **0.01** | **0.03** |
|  | RA | 0.008 | 0.0029 | 3.6(0.18) | 0.05 | 0.13 |
|  | **SLE** | **0.21** | **0.0039** | **3.5(0.013)** | **0.009** | **0.03** |
| *Abbreviations: ALG = allergic disease ; AST = asthma ; Lymph = lymphocyte count ; RA = rheumatoid arthritis ; SLE = systemic lupus erythematosus ; R2 = variance explained by the model (model fit) ; se = standard error; FDR = false discovery rate; bold = signficant association after FDR-correction.* | | | | | | |

Table S 7 Results of regression analyses of polygenic score analyses on autistic-like traits stratified by sex

| Autistic-like traits | Immune phenotype | Males | | | Females | | |
| --- | --- | --- | --- | --- | --- | --- | --- |
|  |  | Beta (se) | p | q | Beta (se) | p | FDR-p |
| Childhood Behavior | Allergy | -0.024  (0.027) | 0.36 | 0.64 | -0.03.5(0.025) | 0.15780 | 0.32 |
|  | Asthma | 0.036 (0.027) | 0.18 | 0.42 | 0.018 (0.025) | 0.46761 | 0.58 |
|  | Lymph | 0.018 (0.027) | 0.50 | 0.72 | 0.071 (0.025) | 0.00469 | 0.09 |
|  | RA | 0.022 (0.027) | 0.40 | 0.64 | 0.0168(0.025) | 0.50792 | 0.60 |
|  | SLE | 0.036 (0.027) | 0.18 | 0.42 | 0.050 (0.025) | 0.04561 | 0.19 |
| Rigidity | Allergy | 0.028 (0.026) | 0.28 | 0.56 | 0.007 (0.025) | 0.7507 | 0.77 |
|  | Asthma | 0.051 (0.026) | 0.04 | 0.37 | 0.039 (0.025) | 0.1156 | 0.26 |
|  | Lymph | 0.012 (0.026) | 0.64 | 0.77 | -0.008 (0.025) | 0.7481 | 0.77 |
|  | RA | 0.039 (0.026) | 0.13 | 0.40 | 0.045 (0.025) | 0.0719 | 0.26 |
|  | SLE | 0.04 (0.026) | 0.06 | 0.37 | 0.040 (0.025) | 0.1107 | 0.26 |
| Social skills | Allergy | -0.05 (0.027). | 0.04 | 0.37 | -0.026 (0.025) | 0.294 | 0.40 |
|  | Asthma | -0.036 (0.027) | 0.18 | 0.42 | 0.030 (0.0252) | 0.226 | 0.36 |
|  | Lymph | 0.028 (0.027) | 0.91 | 0.94 | 0.064 (0.025) | 0.0101 | 0.09 |
|  | RA | -0.056 (0.027) | 0.03 | 0.37 | 0.013 (0.025) | 0.602 | 0.69 |
|  | SLE | -0.011 (0.027) | 0.68 | 0.78 | 0.058 (0.025) | 0.0194 | 0.11 |
| Attention to detail | Allergy | 0.022 (0.027) | 0.40 | 0.64 | 0.033 (0.025) | 0.183 | 0.34 |
|  | Asthma | 0.044 (0.027) | 0.10 | 0.40 | 0.041 (0.024) | 0.103 | 0.26 |
|  | Lymph | 0.033 (0.027) | 0.19 | 0.42 | -0.020 (0.025) | 0.935 | 0.93 |
|  | RA | -0.012 (0.027) | 0.64 | 0.77 | 0.027 (0.025) | 0.274 | 0.39 |
|  | SLE | -0.040 (0.027) | 0.13 | 0.40 | 0.035 (0.025) | 0.160 | 0.32 |
| Imagination | Allergy | 0.018 (0.026) | 0.49 | 0.72 | 0.020 (0.025) | 0.4117 | 0.53 |
|  | Asthma | -0.015 (0.027) | 0.57 | 0.75 | 0.039 (0.025) | 0.1147 | 0.26 |
|  | Lymph | 0.002 (0.027) | 0.94 | 0.94 | 0.029 (0.025) | 0.2464 | 0.36 |
|  | RA | 0.0040 (0.026) | 0.88 | 0.94 | -0.030 (0.025) | 0.2256 | 0.36 |
|  | SLE | -0.041 (0.026) | 0.12 | 0.40 | -0.010(0.025) | 0.6643 | 0.73 |
| Total | Allergy | -0.051 (0.026) | 0.05 | 0.37 | -0.029 (0.024) | 0.240 | 0.36 |
|  | Asthma | 0.015 (0.026) | 0.56 | 0.75 | 0.053 (0.024) | 0.0323 | 0.16 |
|  | Lymph | 0.0089 (0.023) | 0.73 | 0.81 | 0.06.2 (0.02) | 0.0123 | 0.09 |
|  | RA | 0.044 (0.026) | 0.09 | 0.40 | 0.043 (0.024) | 0.0785 | 0.26 |
|  | SLE | 0.022  (0.026) | 0.38 | 0.64 | 0.064 (0.024) | 0.00973 | 0.09 |
| *Abbreviations: ALG = allergic disease ; AST = asthma ; Lymph = lymphocyte count ; RA = rheumatoid arthritis ; SLE = systemic lupus erythematosus ; se = standard error; p = p-values; FDR = false discovery rate.* | | | | | | | |
